# Folic acid prevents interferon-induced iron accumulation and ferroptosis and improves liver health in children with biliary atresia

**DOI:** 10.1101/2023.12.01.23299102

**Authors:** Yanhui Xu, Xixi Chen, Rongli Fang, Yu Ning, Zhijun Zhu, Xiaolei Wang, Yunnan Xiao, Xiaotian Li, Huifang Ren, Yanfang Zhang, Xiaoyu Zuo, Chengwei Chai, Kanghua Zhong, Jiankun Liang, Qifeng Liang, Yuanyuan Luo, Yi Xu, Kaili Liao, Qiuming He, Xuying Tan, Qingqing Ye, Zefeng Lin, Yang Han, Zhenhua Luo, Xiaoqiong Gu, Yan Zhang, Liying Sun, Fan Bai, Jinbao Liu, Junqiang Lv, Zhi Yao, Andrew M Lew, Huimin Xia, Wenhao Zhou, Zhe Wen, Zhanghua Chen, Yuxia Zhang

## Abstract

**Background & Aims:** Biliary atresia (BA) is an obstructive newborn jaundice disease that leads to liver failure in the majority of affected infants. Viral infection is an important environmental trigger of BA. The aim of the study is to establish how viral infection rewires the cellular and metabolic processes of the digestive systems in at-risk infants and leads to BA development.

**Methods:** Single cell RNA (scRNA) transcriptomes and V(D)J sequences were generated using small intestine and liver biopsies from BA and control infants. Candidate risk genes were identified by genome-wide association study. Patient specimens, mouse model of experimental biliary atresia, and a myeloid-specific *Folr2* knockout mice (*folr2^Mko^*) were used to determine immune pathologies that lead to BA development. An open label clinical trial was conducted to determine the therapeutic effect of folic acid on post-Kasai’s outcomes of patients with BA.

**Results:** Type I interferon (IFN-I) signaling is persistently activated in infants with BA. This promotes expression of hepcidin in hepatic TREM2^+^ macrophages and hepatocytes, which impairs SLC40A1-mediated iron excretion from the small intestine, leading to iron accumulation, lipid peroxidation, dysbiosis and folic acid deficiency. By genetic ablation of *Folr2,* we show that folinate supplementation halts persistent IFN-I activation and suppresses hepcidin expression by TREM2^+^ macrophages. In an open label clinical study, folic acid supplementation decreased post-Kasai’s cholangitis incidences and liver transplantation rates by 70%.

**Conclusion:** Persistent IFN-I signaling plays a critical role in virally induced pathological jaundice in infants, and that folic acid supplementation is an effective therapy for BA.

## INTRODUCTION

Bilirubin and iron are metabolic products of heme, which are generated by macrophage processing of senescent or damaged erythrocytes. Most circulating iron in blood plasma is recycled and transported to the bone marrow for erythropoiesis ^1^. Unconjugated bilirubin (UCB) is lipophilic and when in excess may accumulate in the brain and cause life-threatening kernicterus in neonates. In plasma, UCB is bound by albumin and delivered to the liver for glucuronidation by UDP-glucuronosyltransferases (UGT, encoded by *UGT1A1*). This generates water soluble conjugated bilirubin (CB), which is subsequently transported to the bile canaliculi via MRP2 (encoded by *ABCC2*). CB, bile salts and acids, are stored in the gallbladder and released into the small intestine upon food ingestion ^2^.

In the neonates, physiological jaundice is a benign condition with transient elevation of blood bilirubin concentrations. It occurs because neonatal erythrocytes are prone to oxidative stress and have a shortened life span of 60-90 days compared to that of adult erythrocytes of 120 days ^3^. Pathological jaundice, in contrast, is a multifactorial disorder that involves excessive breakdown of erythrocytes, insufficient UCB glucuronidation, impaired CB excretion or bile duct obstruction. G6PD deficiency ^4^, ABO hemolytic disease of the newborn (ABO HDN) ^5^ and viral infection ^6^ are common triggers of neonatal pathological jaundice with variable degrees of clinical severity. Biliary atresia (BA) is an obstructive pathological jaundice disease that affects one in 5,000-18,000 newborns ^7^. Hepatoportoenterostomy (Kasai’s procedure) is usually performed to remove bile duct remnants and re-connect the bile flow. Despite this reconstruction, intestinal dysbiosis, pathobiont accumulation, and bacterial cholangitis develop frequently in BA ^8^. As a result, most infants with BA are destined for jaundice recurrence and require liver transplantation ^7^.

Although some infants with BA display major congenital extrahepatic anomalies ^9^, the disease is commonly regarded as non-Mendelian and resembles a complex trait ^10^, with obliterative extrahepatic cholangiopathy as the common endpoint. The proposed mechanisms include environmental triggers, abnormal immune responses, and genetic predispositions ^11^. Of particular interest, hepatotropic viruses are detected in 28-42% BA patients by polymerase chain reaction (PCR) at the time of Kasai’s procedure ^12–14^. Mx-A (encoded by *MX1* gene), an IFN-I induced gene that is indicative of previous or active viral infection, is positive in 92% of the liver biopsies by immunohistochemistry ^15^. Consistent with viral infection being a candidate trigger for BA, Rhesus rotavirus (RRV) infection induces extrahepatic bile duct obstructions in the neonatal BALB/c mice ^16, 17^. Therefore, promoting early antiviral activity by infusion of a recombinant IFN-α ^18^, or by blocking virally-induced activation of T helper 1 (Th1) cells ^19^ or B cells ^20, 21^, decrease BA incidence in RRV-infected neonatal mice.

Despite that viral infection represents an attractive environmental etiology for BA susceptibility, in human neonates, the cellular and molecular mechanisms linking viral infection to BA remain unresolved. Here, we performed scRNA transcriptome profiling using small intestine and liver biopsies from control subjects and those with BA. We examined the sequential immune and metabolic perturbations (in folic acid and iron pathways) using the RRV-induced BA model. We showed that persistent IFN-I signaling (such as that induced by viral infections) in at-risk individuals, plays a critical role in BA pathogenesis and that folic acid supplementation is a promising therapy.

## METHODS

### Ethical statement

Study procedures were approved by the Medical Ethics Committee of Guangzhou Women and Children’s Medical Center (GWCMC) (ID: [2021]441B00). The implementations were in concordance with the International Ethical Guidelines for Research Involving Human Subjects as stated in the Helsinki Declaration. The legal guardians of all participants signed informed consent forms for biological investigations.

### Demographic characteristics of participants

For laboratory experiments, we established a cohort of patients from the Department of Pediatric Surgery at GWCMC who were enrolled between July 2020 and June 2023. BA (n =126) (median age: 2 months, ranging from 18 days to 5 months; male/female = 1.13) was diagnosed when intrahepatic biliary tree and/or extrahepatic bile duct could not be observed with intraoperative cholangiography. For controls, children with normal liver function but requiring surgery for choledochal cyst (CC) (n = 152) (median age: 24 months, ranging from 5 days to 5 years 8 months; male/female = 0.74) were included. Small intestine and liver biopsies from BA (n=18) and control-CC infants (n=13) were obtained for scRNAseq. Folic acid supplementation was assigned randomly to 19 patients with BA. Epidemiological data and information for postoperative autologous liver survival were summarized in Supplementary table 1. In the cohort study, the population under investigation consists of individuals who were diagnosed with CC (n=17), G6PD deficiency (n=27), ABO HDN (n=30), HCMV^+^NH (n=62) or BA (n=66). Infants were eligible for inclusion if: Full term neonates; Signed informed consent was obtained. Infants were not eligible if: The infant received phototherapy or exchange transfusion; age over 5-month-old. The inclusion and exclusion criteria were also adapted to selection of age-matched healthy controls. HCMV/RV-specific antibodies, iron contents and FA concentrations were measured on the remaining blood samples after clinical laboratory testing. Epidemiological data of this cohort was summarized in Supplementary table 1.

### Clinical survey

The survey, conducted by Department of Pediatric Surgery at GWCMC, obtained information using electronic forms. The survey included 249 valid cases in total, 154 from patients with BA, 95 from patients with CC. Participants were recruited via online follow-up applet (compatible for both computer and hand-held devices). Participants completed the online survey upon invitation. The main contents of the structured questionnaire included nutritional supplementations of the mothers during pregnancy and after childbirth, postnatal nutrition and infection and postoperative complications of the infants (Supplementary table 2). The survey has acquired approval from the Medical Ethics Committee of GWCMC for data collection and analysis.

### Genome-wide association study (GWAS)

Peripheral blood from 297 patients with BA from GWCMC and Liver Transplantation Center, Beijing Friendship Hospital were collected as a case cohort for the GWAS study. The legal guardians of all participants signed informed consent forms for biological investigations. This project was reviewed and approved by the Ethics Committee of GWCMC and Ethics Committee of Beijing Friendship Hospital, in adherence with the Declaration of Helsinki Principles. After quality control, 5,955,093 genotyped or imputed SNPs from 281 cases and 3453 controls were obtained in this study. PLINK (v1.9b) program was used for quality control ^22^. Whole-genome imputation of SNPs was carried out by using the IMPUTE2 program ^23^. Gene-based association analyses were conducted by using the effective chi-squared test (ECS) method implemented in KGG (v4.0) software ^24^.

### Clinical trial

For assessing the therapeutic effects of folic acid supplementation, an open label clinical study was conducted in 19 patients (average days at surgery: 68 days, ranging from 44 days to 82 days) with BA (Chinese clinical trial, ChiCTR2100050992) between 15 September 2021 and 22 February 2022 at GWCMC, Guangzhou, China. Patients enrolled received orally folic acid 0.4mg, once daily. After 3-7 days, patients received Kasai’s procedure. The postoperative course involved nasogastric drainage, intravenous fluids and vitamins supplement until there was evidence of bowel function. Folic acid tablet was added after the infants began normal feeding (0.4mg/day), in combination with Ursodeoxycholic acid (10-15 mg/kg bid). Methylprednisolone injection was given 5^th^∼7^th^ days PO at 4 mg/kg qd followed by 2 mg/kg qd, and replaced with tablets 2 mg/kg qd for 3 weeks. Cefoperazone and metronidazole were administered for 10-14 days PO. Cefixime and TMPco were alternately orally taken for 6 months. Patients attended subsequent outpatient visits or received telephone follow-up monthly after operation. Renal function and adverse events were monitored before, during and after folic acid supplementation.

### Biopsies and stool sample processing

Liver and small intestine biopsies were obtained from patients during laparoscopy or Kasai’s operation. The specimens were collected in MACS® Tissue Storage Solution (Miltenyi Biotec) and transferred to laboratory on ice. Each sample was fixed in 4% PFA for histological assessment and the sparing tissue was used for digestion to harvest single-cell suspension. Processing for liver ^21^ and small intestine ^25^ has been described previously. Briefly, hepatic tissues (0.05∼0.2g) were minced, filtered, and collected in PBS with 2% FBS, followed by low-speed centrifuge to remove majority of hepatocytes. 35% Percol buffer was used to enrich immune cells for flow cytometry analysis. Small intestine tissues (0.5*0.5*0.5cm) was cut into small pieces and digested by collagenase 1A (1 mg/mL) and DNase I (10 U/mL) with gentle rotation at 37°C for 30 min. After filtering and washing, single-cell suspensions were used for scRNAseq or phenotypic flow cytometry assays. Stool samples at admission from patients with BA and CC were collected in a clean, dry screw-top container by the nurses in the ward. Stool samples from age-matched healthy controls were collected at clinics of Neonatal Health Department. Stool samples were divided and transferred to −80°C refrigerator for storage immediately. The aliquots were used for measurement of iron and folic acid concentrations.

### Mouse experiments

The animal studies were approved by the Institutional Animal Care and Use Committee of Guangzhou Medical University (ID: GY2021-119). BALB/c mice were purchased from Laboratory Animal Center of Guangdong Province, China. Breeding strategies were as described ^21^. Briefly, breeding pairs (1 male and 2 or 3 female mice) were housed in microisolator cages under standard laboratory conditions (room temperature 22°C ± 2°C, relative humidity 50% ± 10%) and a 12h/12h light/dark cycle. Once the vaginal plug is observed in females, they were placed into separate housings. Litters of newborn pups (within 12 hours) were assigned to the following groups: (1) control group, which received an intraperitoneal injection with PBS, (2) RRV group, which were intraperitoneally infected with 50 uL PBS containing 1.5×10^6^ plaque forming units (PFUs) of RRV, (3) RRV+ FeSO4•7H2O, 5mg/kg FeSO4•7H2O (Sigma) was gavaged to RRV-infected mice every day, (4) RRV+anti-IFNAR-6hrs, 6 hours before RRV infection, anti-IFNAR1 (Biocompare, 50mg/kg, Clone: MARI-5A3, mouse IgG1κ) was given intraperitoneally and re-supplemented at the 6^th^ day post RRV infection, (5) RRV+anti-IFNAR-18hrs, same dosage of anti-IFNAR was given 18 hours after RRV infection and re-supplemented at the 6^th^ day post RRV infection, (6) RRV+JAKi, Tofacitinib (Selleck, 15mg/kg) was administered 18 hours post RRV infection and daily injected. (7) RRV+CF (Calcium folinate, MCE), first dose of CF (0.5mg/kg) was injected 18 hours after RRV infection and continued daily until sacrifice, (8) RRV+rIFNβ-transient, recombinant human IFN-β (Peprotech, 10,000U) was injected 6 hours after RRV infection and re-supplemented at the 6^th^ day post RRV infection, (9) RRV+rIFNβ-persistent, recombinant human IFN-β (Peprotech, 10000U) was injected 18 hours post RRV infection and re-supplemented every day until sacrifice. (10) RRV+Vitamin E (VitE, Selleck), first dose of VitE (40mg/kg) was injected 18 hours after RRV infection and given every two days until sacrifice. All protocols were approved by Animal Ethics Committee of Guangzhou Medical University, Guangzhou, China. Mice were monitored daily and euthanized on the 12^th^ or 13^th^ day.

### Generation of Folr2^flox/flox^Lyz2-Cre^+^ (Folr2 ^Mko^) mice

The loxP sequences were introduced by CRISPR/Cas9 technology to allow for conditional deletion of exon 4-6 of *Folr2*, which would result in a null allele upon Cre recombinase-mediated excision. In order to get *Folr2*-floxed offspring on BALB/C background, *Folr2^flox/flox^* mice on C57BL/6N background were intercrossed with wild type BALB/C mice for at least six generations. *Folr2^flox/+^* mice from each generation were selected to mate with wild type BALB/C mice. *Folr2^flox/flox^* mice were obtained from the offspring of male and female mice with the *Folr2^flox/+^* gene on BALB/C background. BALB/C-Lyz2em1(Cre-IRES-EGFP)^Smoc^ mice (purchased from Shanghai Model Organisms Center Inc.) were intercrossed with *Folr2^flox/flox^*mice to obtain *Folr2^flox/+^Lyz2-Cre^+^* mice. When enough pregnant mice were obtained, litters of their newborn pups were assigned to the following groups: (1) control group, which received an intraperitoneal injection with PBS, (2) RRV group, which were intraperitoneally infected with 50 uL PBS containing 1.5×10^6^ plaque forming units (PFUs) of RRV within the first neonatal 12 hours, (3) RRV+CF (Calcium folinate, MCE), first dose of CF (0.5mg/kg) was injected 18 hours after RRV infection and continued daily until sacrifice. The genotypes were identified by polymerase chain reaction (PCR). Sequences of forward and reverse primers are listed in Supplementary table 3.

### RNA-seq library construction for 10x Genomics single cell 5’ and V(D)J sequencing

Liver and small intestine biopsies were processed as described ^21, 25^. For liver, CD45^+^ live cells were flow-sorted by a FACSAria SORP flow cytometer (BD Sciences). For small intestine, single cell suspensions were obtained after enzymatic digestion and dead cell removal (Miltenyi Biotec). Single-cell 5’ RNA sequencing libraries were constructed according to the protocols of the Chromium Single Cell 5’ Library kit (10x GENOMICS). In brief, single-cell suspension was mixed with RT-PCR master mix and loaded onto the 5’ chip. Then, RNA transcripts were uniquely barcoded within each cell and reverse-transcribed into barcoded cDNA, followed by purification, amplification and adaptor ligation. TCR- and BCR-enriched libraries were generated with aliquots from each of the aforementioned cDNA using the Chromium Single Cell V(D)J Enrichment kit. All libraries were sequenced on the Illumina Novaseq 6000 platform.

### RNA-seq library construction for BD Rhapsody™ single-cell analysis system

Liver and small intestine biopsies from age matched infants (8 BA, 4 age-matched control, 5 BA with FA treatment) were processed as described above. BD Rhapsody^TM^ Human Single-Cell Multiplexing Kit (BD biosciences) was utilized for scRNA library construction. Up to 2 samples were labelled and pooled prior to single cell capture with the BD Rhapsody™ Single-Cell Analysis system. After partitioning and lysis of cells, cDNA is encoded on BD Rhapsody™ Enhanced Cell Capture beads using both the 3’ and 5’ ends of transcripts as templates. Whole transcriptome mRNA libraries are amplified using random priming of the on-bead cDNA libraries.

### Preprocessing of single-cell RNA-seq data

The raw sequence reads were demultiplexed and aligned to the human transcriptome reference (GRCh38) using CellRanger-3.1.0 (for 10X Genomics) or a cwl pipeline (for BD Rhapsody^TM^ system) (https://bitbucket.org/CRSwDev/cwl/src/master/). The raw expression count matrix (gene counts versus cells) was generated for each sample and transformed to Seurat object using the R package Seurat v4.0.0 ^26^. Qualified cells are identified as cells with 200-5000 detected genes and with less than 50,000 detected unique molecular identifiers (UMIs). Following quality control, the filtered data objects of all samples were merged and normalized using the Seurat’s SCTransform function to eliminate the impacts of sequencing depth, mitochondrial gene expression and other batch effects. A total of 242,327 high quality cells generated using 10X Genomics platform and 210,661 cells generated using BD Rhapsody^TM^ system were included in subsequent analysis.

### Statistical analysis

R v4.0.3 (Foundation for Statistical Computing) was applied for the statistical analyses and graphics production with scRNA-seq data. For the experimental data, GraphPad Prism 9 (GraphPad Software) was used to perform statistical analyses and graphics production. Comparison for categorical variables was conducted using Fisher’s exact test or Chi-squared test. Comparison for continuous variables was conducted using Student’s t test or Mann-Whitney U test. Comparison for different factors (folic acid treatment, surgeons, gender and age at surgery) on the clinical results between the groups of BA with or without folic acid-treatment was conducted using logistic regression model. The difference of cell proportions between different groups based on single-cell sequencing data was also verified by using a Poisson regression model as previously described ^27^. The P value for the significance was assessed by the Wald test on the regression coefficient. All the statistical analyses were performed in two-sided manner. Detailed descriptions of other specific statistical tests are specified in the relevant Results section and Figure Legends.

## RESULTS

### Establishing the small intestine and liver cellular landscape for control infants and those with BA

To relate viral infection to BA pathogenesis, we first performed a questionnaire survey on control subjects and infants with BA. Control subjects were infants who had normal liver function but required surgical interventions due to choledochal cysts (CC, hereafter labelled control-CC) ^21^. The survey showed that prenatal maternal folic acid supplementations were not significantly different between the two groups (Supplementary Figure 1A). However, human cytomegalovirus (HCMV) infection and post operational bacterial cholangitis occurred significantly more frequently in BA than control-CC subjects (Supplementary Figure 1A). Of this survey cohort, 39.7% of patients with BA underwent liver transplantation. For those with liver transplantation, 87% of patients received this surgery within 1.5 years of age (Supplementary Figure 1A).

To further determine involvement of viral infection in BA, we compared serum viral-specific humoral responses from an independent cohort of healthy control subjects (hereafter labelled control-healthy), infants with BA or infants with a diagnosis of HCMV-positive neonatal hepatitis (HCMV^+^ NH). HCMV-specific IgG and IgA antibodies, as well as human rotavirus (RV)-specific IgG and IgM antibodies were significantly increased in BA and HCMV^+^ NH infants compared with the control-healthy subjects (Supplementary Figure 1B). Immunofluorescent staining confirmed the serological findings and showed that HCMV and RV antigens were significantly enriched in BA compared with control-CC infants. Viral antigens were identified in the liver hepatic cells, vascular endothelial cells, bile duct epithelial cells and inflammatory infiltrating cells (Supplementary Figure 1C).

To define how viral infection may rewire the intestinal and hepatic cellular compositions and interactions, we then generated single cell RNA (scRNA) transcriptomes and V(D)J sequences using small intestine and liver biopsies from BA and control-CC infants (Figure 1A; Supplementary Figure 2A-B). After data pre-processing and quality control, we identified 7 major cell clusters for the small intestine and 5 major cell clusters for the liver. All cellular subsets had representative cells from each individual sample; batch effects were negligible (Supplementary Figure 2C-E).

**Figure 1.**
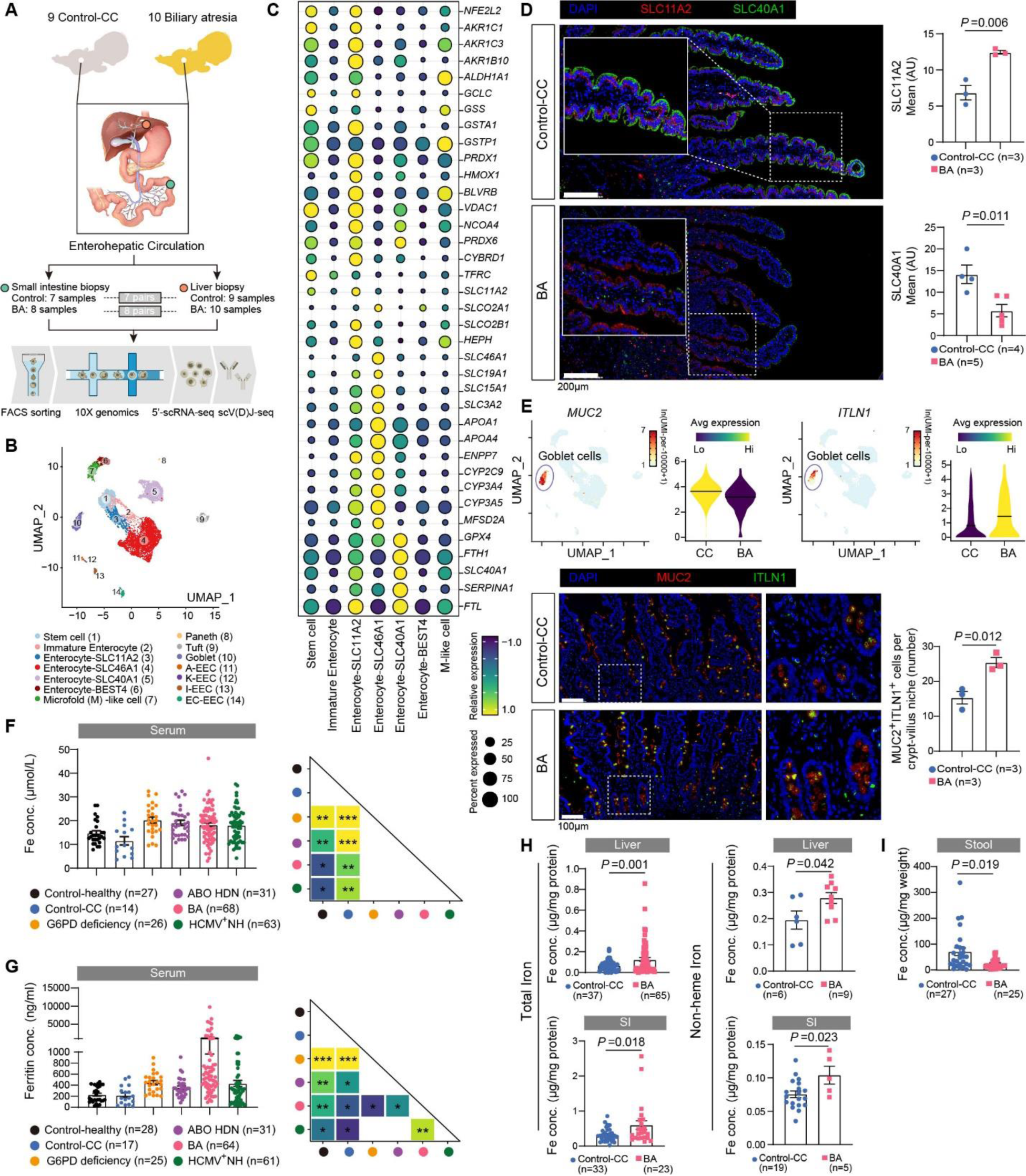
Infants with BA Display Systemic Iron Accumulation and Luminal Iron Deficiency. (**A**) Schematic diagram showing experimental design for 10x Genomics single cell 5’- and V(D)J-sequencing (scRNA-seq). (**B**) UMAP plot displaying distributions of 14 subsets of cells that constitute the small intestine epithelium. (**C**) Dot plots displaying expression of genes mediating heme and bilirubin metabolism, glutathione biosynthesis, folic acid and fatty acid transportation, lipid oxidation as well as iron storage and exportation across different enterocyte clusters. (**D**) Representative immunofluorescent images showing distribution of SLC40A1 (green) and SLC11A2 (red) in small intestine. Scale bar, 200μm. Measurement of mean fluorescence intensity (MFI) for SLC40A1 and SLC11A2 and the statistical unit was shown as Arbitrary Unit (AU). (**E**) UMAP plots showing expressions of MUC2 and ITLN1 in small intestinal enterocytes. Color intensity is proportional to normalized unique molecular identifier (UMI) counts in each cell. Violin plots showing relative expression of MUC2 and ITLN1 in goblet cells. Representative immunofluorescent images showing expressions of MUC2 (red) and ITLN1 (green) in small intestine. Scale bar, 100μm. Average numbers of MUC2^+^ITLN1^+^ cells per crypt-villus from BA (n=3) and control (n=3) subjects were counted. (**F** and **G**) Dot plots showing concentrations of serum iron and ferritin in age matched control-healthy (n=27), control-CC (n=14), G6PD deficiency (n=25), ABO HDN (n=31), BA (n=64) and HCMV^+^NH (n=61). Pair-wised statistical comparison results were shown, and color represents the P value. Student’s t test, *P < 0.05, **P < 0.01. (**H**) Dot plots showing concentrations of total and non-heme iron in homogenates of liver and small intestine biopsies. (**I**) Dot plots showing iron content in stool, shown as per mg weight. For all data, each point represents an individual patient, and the line represents the mean ±SEM. *P* values were calculated by two-tailed Student’s t test unless otherwise indicated.

### Infants with BA display systemic iron accumulation and luminal iron deficiency

We first examined small intestinal epithelial cells (Figure 1B; Figure S3A to S3C). In support of the physiological importance of enterohepatic circulation in neonates, we found that genes that regulate heme and bilirubin metabolism as well as iron storage and export were highly and differentially expressed by the small intestinal enterocytes (Figure 1C). Specifically, we found that enterocytes-SLC46A1 cells expressed genes for heme transport and breakdown (*SLC46A1*, *CYP2C9*, *CYP3A4*, *CYP3A5*) and absorption of bile acid (*SLC15A1*), folic acid (*SLC46A1*, *SLC19A1*) and fatty acid (*APOA1*, *APOA4*, *ENPP7*). Enterocytes-SLC11A2 cells expressed genes for iron absorption (*SLC11A2*, *HEPH*, *TFRC*), bilirubin metabolism (*SLCO2B1*, *HMOX1*, *BLVRB*), glutathione biosynthesis (*GCLC*, *GSTA1*) and anti-oxidative stress responses (*NFE2L2* encodes NRF2, *GPX4*). Enterocytes-SLC40A1 cells export iron across epithelia and were enriched with intracellular iron storage proteins (*FTH1* and *FTL*) and *GPX4* (Figure 1C).

To determine whether iron metabolism contributes to BA pathogenesis, we examined the expression of iron transporters. In the small intestinal epithelial cells of age-matched control-CC infants, SLC11A2 (also known as DMT1, iron absorption) was strongly expressed at the basolateral membrane, while SLC40A1 (also known as FPN1, iron exportation) was mainly located at the apical (luminal) membrane (Figure 1D; Supplementary Figure 3D). In control-CC infants, we surmise that this spatial polarity may allow export of excessive iron from the circulation to the intestinal lumen, which may serve as a protective mechanism from the excessive erythrocyte damage in neonates ^3^. However, in infants with BA, this iron transfer may be impaired as much of the SLC11A2 was now located on the luminal membrane and levels of SLC40A1 were significantly decreased (Figure 1D). Lactoferrin is the major dietary iron in early infancy, we also found that its receptor *ITLN1* was highly increased in goblet cells in infants with BA (Figure 1E). To confirm the above changes to iron status, we measured iron concentrations in the serum and the hepatic and small intestinal biopsies from control-CC and BA infants. We also included serum samples from children with other types of cholestatic diseases, including G6PD deficiency, ABO HDN, and HCMV^+^ NH. While serum iron concentrations were significantly elevated in infants with cholestatic diseases (Figure 1F), serum ferritin levels, which reflects the amount of iron stored in the body, were most highly elevated in BA compared with control subjects and children with other types of cholestatic diseases (Fig. 1G). It should be noted that the increased plasma iron concentration and decreased intestinal expression of *Slc40a1* were also observed in the bile duct ligation (BDL) model and in H1N1 influenza (PR8) infected suckling mice (Supplementary Figure 3E-F). Hepcidin, the liver-derived peptide hormone that suppresses SLC40A1-mediated iron excretion ^28^, was significantly increased in BDL and H1N1 influenza infected mice compared with control mice (Supplementary Figure 3E-F). We thus envisage that iron accumulation is common in neonatal cholestatic diseases caused by excessive erythrocyte damage (such as G6PD deficiency, ABO HDN), bile duct obstruction (such as in the BDL mouse model), and by viral infection (HCMV^+^ NH, BA, H1N1 influenza infection).

Direct measurement of tissue iron and non-heme iron concentrations confirmed significant elevation in BA compared with control-CC subjects (Figure 1H). We also measured iron from the stool samples. Infants with BA showed significantly decreased luminal iron concentrations compared with age-matched control-CC subjects (Figure 1I). These differential iron levels and the SLC40A1 findings would be concordant with the view that systemic iron accumulation was a consequence of impaired luminal iron excretion. Collectively, we show that infants with BA display abnormal expression and distribution of the iron transporters in the small intestine, resulting in systemic iron accumulation and luminal iron deficiency.

### Redox imbalance promotes iron-induced lipid peroxidation and ferroptosis of the small intestine and liver

In order to identify mechanisms that lead to systemic iron accumulation, we examined differentially expressed genes of the small intestinal enterocytes between BA and control subjects. Genes that regulate response to IFN-I and IFN-γ, activation of JAK-STAT cascade, iron transport and microvillus organization were significantly enriched in BA compared with control-CC subjects (Figure 2A; Supplementary Figure 4A). We measured serum IFN-I and found that IFN-β was significantly increased in BA compared with infants with non-viral cholestatic diseases (Figure 2B). Additionally, we found that genes that inhibit lipid peroxidation (*SLC40A1*, *GPX4*, *SELENOP*) were significantly decreased in BA compared with control-CC subjects (Supplementary Figure 4A). Immunofluorescent staining of GPX4 and its upstream regulator NRF2 confirmed that their protein expressions were also significantly decreased in the small intestine and liver in BA compared with control-CC subjects (Supplementary Figure 4B).

**Figure 2.**
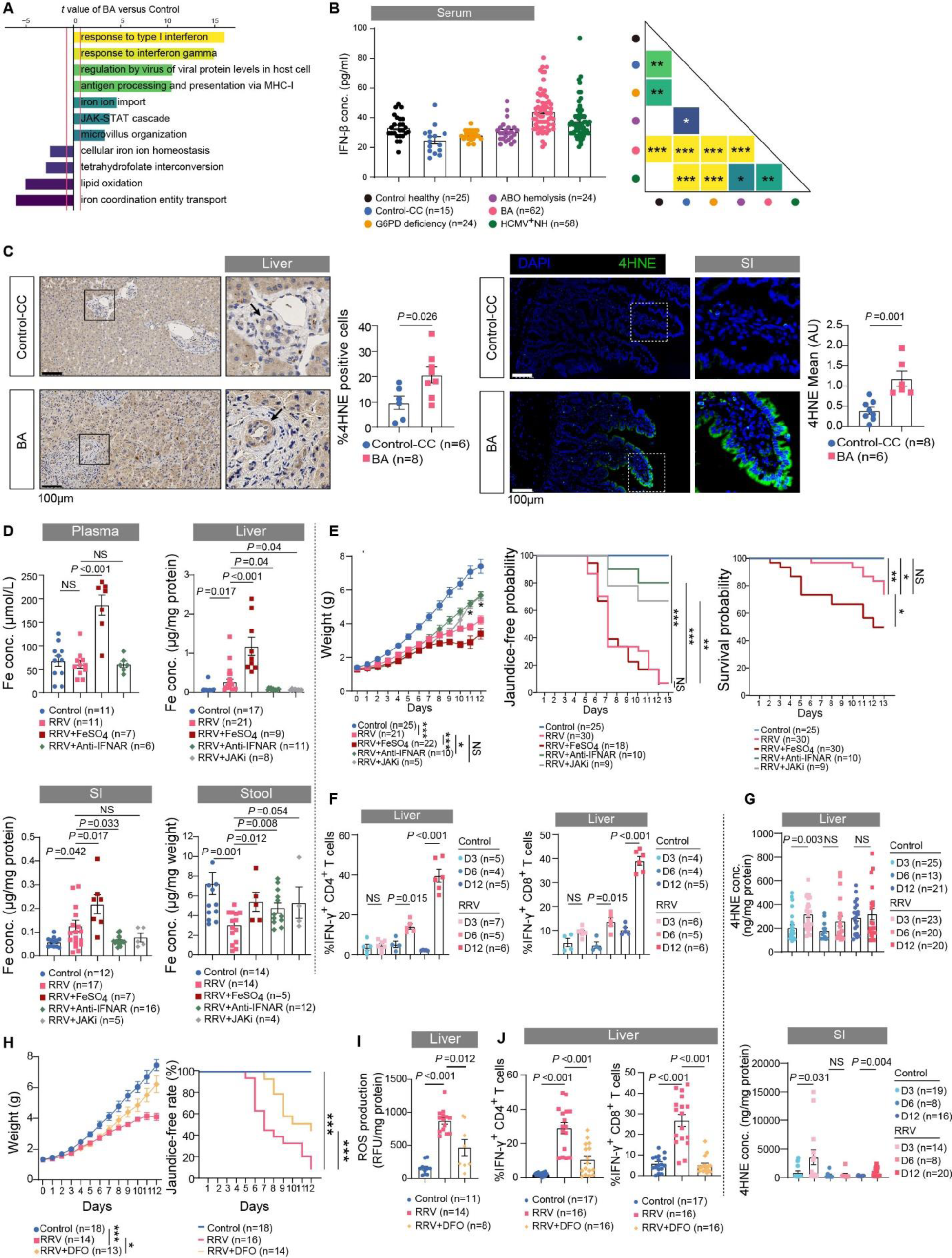
Interferon-iron Imbalance Induces Lipid Peroxidation and Ferroptosis of the Small Intestine and Liver. (**A)** Bar plots showing enriched GO terms of the genes overexpressed by enterocyte-SLC11A2, enterocyte-SLC40A1 and enterocyte-SLC46A1 subsets between BA and control-CC subjects. The average *t* statistic across cells was determined by GSVA analysis for two groups. (**B)** Serum levels of IFN-β in age matched infants with control-healthy (n=25), control-CC (n=15), G6PD deficiency (n=24), ABO HDN (n=24), BA (n=62) and HCMV^+^NH (n=58). Pair-wised statistical comparison results were shown, and color represents the P value. Student’s t test, *P < 0.05, **P < 0.01, ***P < 0.001. (**C**) Representative immunohistochemistry images showing 4-HNE expression in liver biopsies. Black arrow points to biliary epithelial cells (BECs). Quantifications are percentage of 4-HNE^+^ cells per mm^2^ visual field. Scale bar, 100μm. Representative immunofluorescent images showing 4-HNE staining in the small intestine biopsies. Scale bar, 100μm. (**D**) Dot plots showing concentrations of iron in plasma, stool, and homogenates of liver and small intestine. (**E**) Line charts displaying body weight changes (left), jaundice occurrence rates (middle) and survival probabilities (right). The statistics for body weight change was performed using two-way ANOVA, for jaundice-free survival rate and survival probability were performed using Log-rank test; *p < 0.05, **p < 0.01, ***p < 0.001. (**F**) Dot plots showing the frequencies of intrahepatic IFN-γ^+^CD4 T and IFN-γ^+^CD8 T cells measured by flow cytometry. (**G**) Dot plots showing the concentrations of 4-HNE (in the small intestine and liver) from control and RRV-infected mice at indicated time. (**H**) Line charts displaying body weight changes and jaundice occurrence rates. The statistics for body weight change and jaundice-free survival were performed using Two-Way ANOVA and Log-Rank test, respectively; *p < 0.05, **p < 0.01, ***p < 0.001. (**I**) Dot plots showing hepatic ROS production(**J**) Dot plots showing the frequencies of intrahepatic IFN-γ^+^CD4 T and IFN-γ^+^CD8 T cells. For all data, each point represents an individual patient/mouse, and the line represents the mean ± SEM. *P* values were calculated by two-tailed Student’s t test unless otherwise indicated.

GPX4 is a critical negative regulators of lipid peroxidation and ferroptosis ^29^. We next determined whether its decrease may promote iron-dependent lipid peroxidation and ferroptosis ^30^. Transmission electron microscopy of the small intestinal enterocytes revealed that the microvilli were shorter and sparser at the brush borders in infants with BA compared with control-CC subjects. Mitochondria of enterocytes also showed anatomical impairment in BA: loss of cristae, irregular matrix with large voids, and membrane ruptures (Supplementary Figure 4C). 4-Hydroxy-2-Nonenal (4-HNE), a highly toxic aldehyde product of lipid peroxidation, was significantly elevated in the small intestine and liver (particularly in bile duct epithelial cells) in BA compared with control-CC subjects (Figure 2C).

In order to determine whether persistent IFN-I activation contributes to BA pathogenesis via iron metabolism, we utilized the Rhesus rotavirus (RRV)-induced BA model in neonatal BALB/c mice ^31^. On cholangiography, typical clinical BA appearance is characterized by extrahepatic bile duct atresia with a small, fibrotic gallbladder without passage into biliary duct (Supplementary Figure 5A). In RRV-induced BA mice model, edematous swelling of the whole extrahepatic bile duct with short or long-segmented atresia can be observed under dissection microscope (Figure S5B). Consistent with our findings in infants with BA, neonatal RRV infection significantly increased liver and intestinal but decreased luminal iron concentrations (Figure 2D).

Dietary iron supplementation further significantly increased iron overload, inhibited neonatal growth and increased mortality in RRV-induced BA mice (Figure 2D and 2E). In contrast, IFN-I or JAK-STAT signaling blockade ameliorated iron accumulation and BA pathology and improved neonatal growth in RRV-infected mice (Figure 2 D and 2E; Supplementary Figure 5C). Representative histological findings, such as steatosis, necrosis foci of liver, vacuolated epithelium and swelling of small intestine that were exacerbated by RRV infection and with iron supplementation, were ameliorated with anti-IFNAR or JAKi (tofacitinib) treatment (Supplementary Figure 5C).

Activation of IFN-γ-expressing T cells has been reported to contribute to BA liver injury ^21^. We pondered whether lipid peroxidation and ferroptosis may precede adaptive immune activation in BA. In a time-course experiment using RRV-induced BA model, we found that inflammatory cell infiltration at the hepatic portal area, ballooning denaturation of the intestinal epithelial cells (Supplementary Figure 5D), and activation of the IFN-γ-producing T cells (Figure 2F), became prominent 6 days following RRV infection. However, lipid peroxidation (as indicated by elevated 4-HNE levels) had already occurred in liver and small intestine of RRV-infected mice at day 3 (Figure 2G), therefore preceding adaptive immune activation. In addition, deferoxamine (DFO)-mediated iron chelation significantly decreased RRV-induced BA incidence, improved neonatal growth and ameliorated liver histological damage (Figure 2H; Supplementary Figure 5C). Liver reactive oxygen species (ROS) production and hepatic T cell IFN-γ expression were also significantly decreased by DFO treatment (Figure 2I and 2J). IFN-I blockade or JAKi also significantly decreased hepatic T cell IFN-γ expression in RRV-infected mice (Supplementary Figure 5E). VitE, which have been reported to prevent ferroptosis, also reversed RRV-induced BA incidence and improved neonatal growth (Supplementary Figure 5F). Taken together, these results suggest that IFN-I signaling promotes systemic iron accumulation, promotes lipid peroxidation and ferroptosis under the context of GPX4 deficiency, and induces adaptive immune activation.

### Interferon-induced hepatic hepcidin expression promotes iron accumulation

IFN-I is activated upon viral infection in many cell types, including myeloid cells. We identified 11 subsets of myeloid cells in the small intestine and liver (Figure 3A to 3C). Plasmacytoid dendritic cells (pDCs), which produce abundant IFN-I, were mainly detected in the liver. Compared with control-CC subjects, pDCs from infants with BA had significantly increased expression of IFN-I induced gene (*MX1*, *ISG15*, *IRF7*, *IRF8* and *IFIT16*) and decreased expression of the DNA exonuclease *PLD4* (Figure 3D). We also found that intestinal M2 like macrophages (FOLR2^+^Mac) have decreased expression of nucleases including *PLD3*, *PLD4* and *RNASET2* (Figure 3D). Mice that are deficient in *Pld3* and *Pld4* develop lethal hepatitis due to overactivation of IFN-I and IFN-γ signaling ^32^. *Rnaset2* deficiency also induces IFN-I associated neuron pathology ^33^. Analysis of gene signatures confirmed that IFN-I signaling was significantly upregulated in myeloid cells in the small intestine and liver in BA compared with control-CC subjects (Figure 3E).

**Figure 3.**
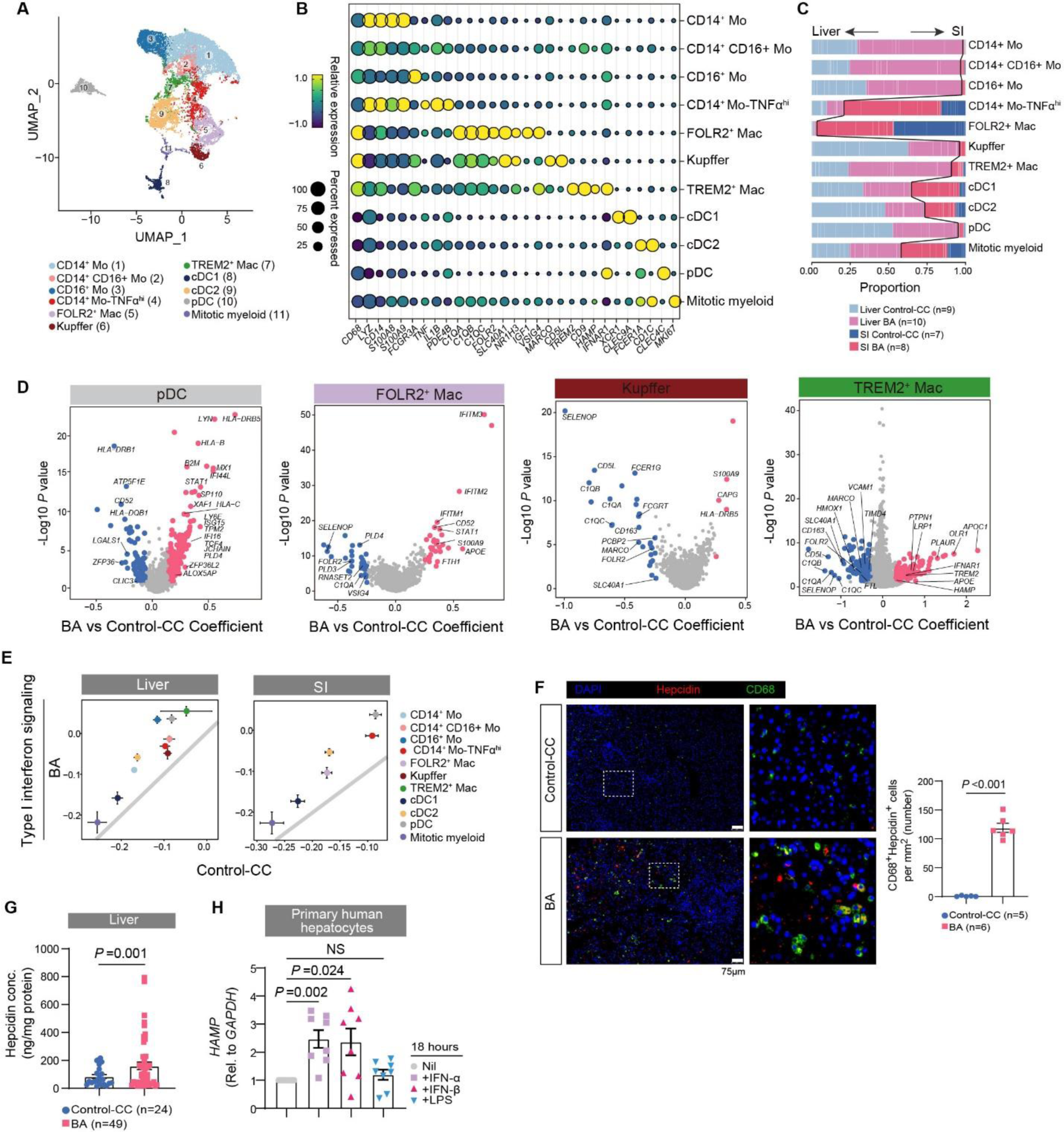
Over-active IFN-I Signaling in Myeloid Cells and Hepcidin Expression in Infants with BA. (**A)** UMAP plots displaying 11 subsets of myeloid cells. (**B)** Dot plots displaying expression of marker genes across different myeloid clusters. (**C**) Bar plots showing cell subset distributions between the different groups. Blocks represent individual samples. Liver and small intestine origins are separated by black line. (**D**) Volcano plots showing differentially expressed genes of pDC, FOLR2^+^Mac, Kupffer and TREM2^+^Mac in comparison to their counterparts from control-CC subjects. Red and blue dots denote the up-regulated and down-regulated genes passing the thresholds of P value <=0.05 and |Coefficient| >=0.25, respectively. (**E**) Scatter plots comparing the signature scores of IFN-I signaling for each subset of myeloid cells in control-CC subjects (x-axis) and infants with BA (y-axis). Dots and lines represent mean ± SEM. (**F**) Representative immunofluorescent images showing co-staining of CD68 (green) and hepcidin (red) in liver. Scale bar, 75μm. Dot plots showing average CD68^+^hepcidin^+^ cell numbers per mm^2^ visual field in the liver biopsies from BA compared with control-CC. (**G**) Dot plots showing concentrations of hepcidin in homogenates of liver. (**H**) qPCR result showing *HAMP* expression in isolated primary hepatocytes under IFN-α, IFN-β and LPS simulation. Each point represents an individual patient, *HAMP* expression was normalized to unstimulated condition (Nil) for each patient. (**I** and **J**) Line charts displaying body weight changes and jaundice occurrence rates. The statistics for body weight change was performed using Two-Way ANOVA, for jaundice-free survival rate and survival probability were performed using Log-Rank test; *p < 0.05, **p < 0.01, ***p < 0.001. (**K**) Relative expression of *Trem2* in liver were examined by qPCR. (**L**) Representative immunofluorescent images showing co-staining of F4/80 (green) and hepcidin (red) in liver. Scale bar, 100μm. Dot plots showing percentage of F4/80^+^hepcidin^+^ cells of F4/80^+^ cells in the liver biopsies. For all data, each point represents an individual patient/mouse, and the line represents the mean ± SEM. *P* values were calculated by two-tailed Student’s t test unless otherwise indicated.

IFNAR expression was found in most myeloid cells, but was highest in pDC and TREM2-expressing macrophages (TREM2^+^Mac) in the liver (Figure 3B). Interestingly, TREM2^+^Mac cells also express *HAMP*, which encodes the iron-regulating peptide hormone hepcidin (Figure 3B). We confirmed that liver macrophages and hepatocytes expressed hepcidin and that hepcidin levels were significantly increased in the liver biopsies in BA compared with control-CC subjects (Figure 3F and 3G). In cultures of primary hepatocytes derived from control-CC subjects, we found that IFN-I (IFN-α and IFN-β) stimulation also directly upregulated expression of *HAMP* in the hepatocytes (Figure 3H).

### Persistent but not transient IFN-I signaling promotes RRV-induced BA pathology in neonatal mice

Although IFN-I has potent anti-viral function, its persistent activation can induce extensive tissue damage ^34^. We therefore set forth to delineate whether transient or persistent IFN-I activation determines BA outcomes in RRV-infected neonatal mice (Supplementary Figure 6A). To reflect transient IFN-I activation, recombinant human IFN-β (rhIFN-β) was injected twice at 6 hours and 6 days post RRV infection ^18, 35^. To maintain continuously activated state of IFN-I sigaling, rhIFN-β was injected daily to RRV-infected neonatal mice. We showed that transient IFN-I activation significantly decreased jaundice incidence (Figure 3I and 3J, labeled as RRV+IFN-β-transient), whereas blocking IFNAR signaling prior to RRV infection provided no therapeutic effects to BA (Figure 3I and 3J). In contrast, persistent IFN-I activation by daily rhIFN-β infusion provided no treatment benefit in RRV-induced BA model (Figure 3J, labelled RRV+IFN-β-persistent), whereas blocking IFNAR signaling 18 hrs post RRV infection significantly decreased jaundice incidence (Figure 3J). Representative histological findings of liver and small intestine were consistent with treatment effectiveness (Supplementary Figure 6B-C).

Mechanistically, we found that RRV infection significantly increased hepcidin expression in hepatic macrophages, and that anti-IFNAR treatment 18 hours post RRV infection abrogated the effects of monocyte infiltration and their differentiation into hepcidin expressing macrophages (TREM2^+^ Mac) at day 12. Transient rhIFN-β infusion did not induce TREM2^+^ Mac expansion (Figure 3K and 3L) and hepcidin elevation (Supplementary Figure 6D).

Finally, we showed that hepatic hepcidin concentrations (Figure 3K; Supplementary Figure 6D) were inversely related with SLC40A1 expression by the small intestine epithelial cells (Supplementary Figure 6E), but positively related with liver iron accumulation and ROS production (Supplementary Figure 6F-G). Therefore, the findings support that persistent IFN-I expression, as seen in infants with BA, promotes BA pathogenesis by promoting hepcidin-iron accumulation and inducing oxidative stress.

### Neonatal viral infection induces intestinal dysbiosis and folic acid malabsorption

Neonatal viral infection decreases iron secretion into the intestinal lumen, which may disturb microbial colonization and maturation in the infant gut ^36^. Therefore, we analysed the gut microbial profile using a published case-control study for BA ^8^. We found that genes that are involved in biosynthesis of antibiotics (vancomycin, streptomycin) and one carbon pool by folate were significantly decreased in the stool samples from BA compared with those from the control subjects (Figure 4A). Furthermore, we measured stool and serum folate concentrations in our own cohort and found that they were significantly decreased in BA compared with age matched control-CC subjects (Figure 4B and 4C). Notably, serum folic acid concentrations were also significantly decreased in HCMV^+^ NH infants (Figure 4C), which suggest that viral infection may impair folate absorption or metabolism. Indeed, folic acid absorption (*Slc46a1*, *Folr2*) and metabolism (*Dhfr*, *Mthfr*) were impaired in RRV-infected neonatal mice (Figure 4D).

**Figure 4.**
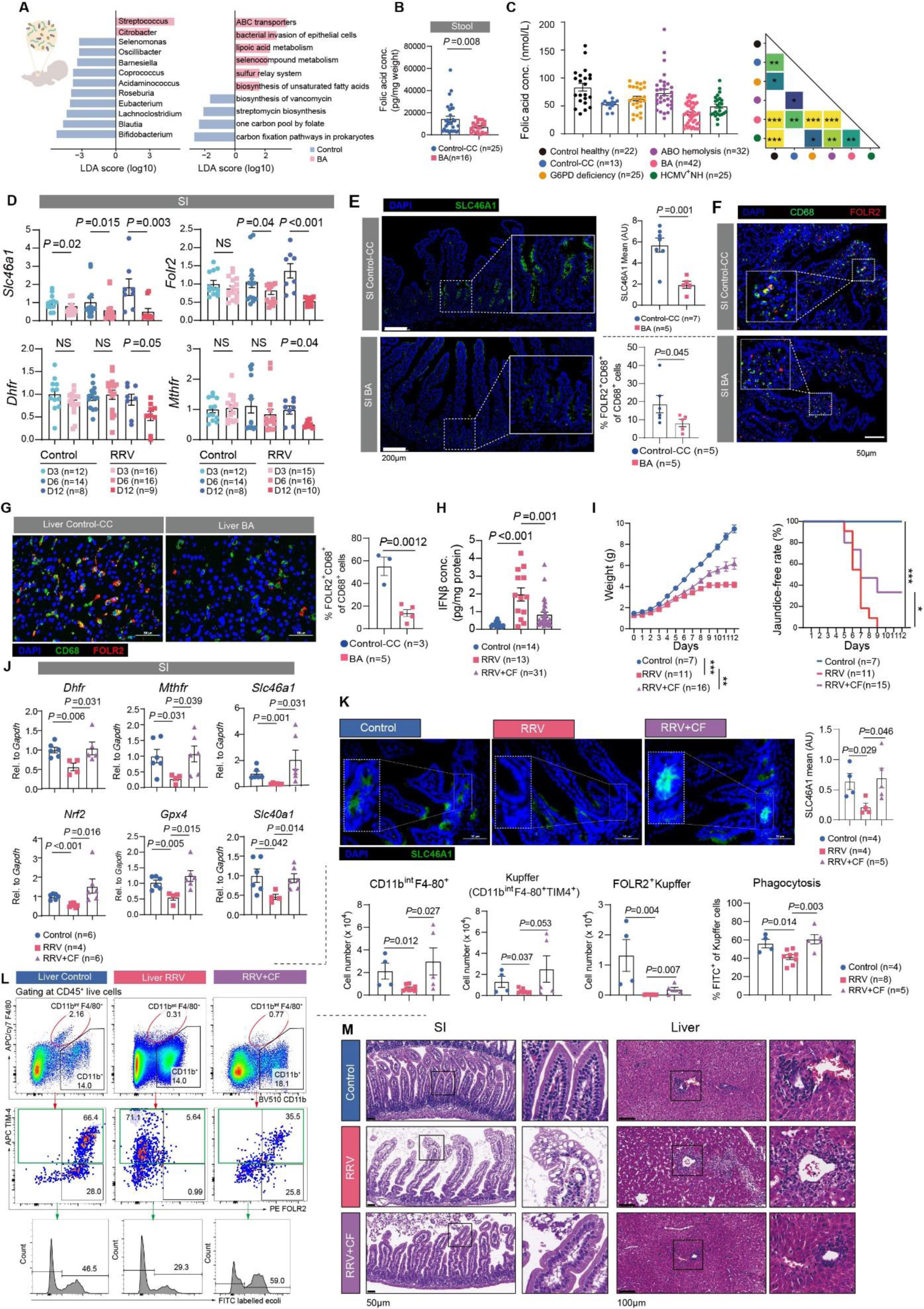
Neonatal Viral Infection Induces Intestinal Dysbiosis and Folic Acid Malabsorption. (**A)** Left panel: bar plots showing LDA scores for the gut microbial abundance at the genus level with significant difference in BA and control subjects. Right panel: Bar plots showing LDA scores for the functional prediction of gut microbial community with significant difference between BA and control subjects, shown as per mg weight. (**B**) Box plots showing the concentrations of folic acid in stool samples. (**C**) Serum levels of folic acid (FA) in age matched infants of control-healthy (n=22), control-CC (n=13), G6PD deficiency (n=25), ABO HDN (n=32), BA (n=42) and HCMV^+^ NH (n=25). Pairwise statistical comparison results were shown, and color represents the *P* value. Student’s t test, *P < 0.05, **P < 0.01, ***p < 0.001. (**D**) Dot plots showing relative gene expression in small intestine from control and RRV-infected mice at indicated time. (**E**) Representative immunofluorescent images showing SLC46A1 staining in the small intestine biopsies. Scale bar, 200um. (**F**) Representative immunofluorescent images showing co-staining of CD68 (green) and FOLR2 (red) in small intestine biopsies. Scale bar, 50μm. Dot plots showing percentage of CD68^+^FOLR2^+^ cells of CD68^+^ cells in small intestine biopsies. (**G**) Representative immunofluorescent images showing co-staining of CD68 (green) and FOLR2 (red) in liver biopsies. Scale bar, 50μm. Dot plots showing percentage of CD68^+^FOLR2^+^ cells of CD68^+^ cells in liver biopsies. (**H**) Dot plots showing concentrations of IFN-β in homogenates of liver from control (n=14), RRV-infected mice (n=13) and with CF treatment (n=31). (**I**) Line charts displaying body weight changes and jaundice occurrence rates. The statistics for body weight change was performed using two-way ANOVA, for jaundice-free survival rate and survival probability were performed using Log-rank test; *p < 0.05, **p < 0.01, ***p < 0.001. (**J**) Dot plots showing relative gene expression in small intestine. (**K**) Representative immunofluorescent images showing SLC46A1 staining in the small intestine biopsies. Scale bar, 50μm. (**L**) Gating scheme for Kupffer cells (CD11b^int^F4/80^+^TIM-4^+^) in the livers. Scavenger function of Kupffer cells was examined by phagocytosis of FITC labelled e-coli. Dot plots showing cell number (*10^4^) of CD11b^int^F4/80^+^ cells, Kupffer cells (CD11b^int^F4/80^+^TIM-4^+^) and FOLR2^+^Kupffer cells from livers. (**M**) Representative Hematoxylin-Eosin (H&E) stained sections for liver and small intestine biopsies. For all data, each point represents an individual patient/mouse, and the line represents the mean ±SEM. *P* values were calculated by two-tailed Student’s t test unless otherwise indicated.

Folic acid supplementation during pregnancy has significantly decreased the incidence of neural tube defects and congenital heart diseases in the human infants ^37, 38^. We found that folic acid transporters were mainly expressed by crypt cells of the small intestine (SLC46A1), as well as by the small intestinal FOLR2^+^Mac (FOLR2^+^CD68^+^) and hepatic FOLR2-expressing Kupffer cells (FOLR2^+^CD68^+^). Both transporters were significantly decreased in infants with BA compared with control-CC subjects (Figure 4E-G).

To determine whether folate supplementation might be useful therapeutically, we intraperitoneally administered calcium folinate (CF, an active form of folic acid that bypasses the impaired folic acid metabolic processes) to RRV-induced BA mice. Remarkably, CF decreased IFN-β concentrations (Figure 4H), improved neonatal growth and decreased jaundice incidence (Figure 4I). Further analysis indicated that genes that regulate folate transportation (*Slc46a1*), metabolism (*Dhfr*, *Mthfr*), oxidative stress (*Nrf2*, *Gpx4*) and iron homeostasis (*Slc40a1*) were all restored in the small intestine of RRV-infected mice that were treated with CF (Figure 4J). Immunofluorescence confirmed that CF treatment increased SLC46A1 expression in crypt cells (Figure 4K). Besides, decreased FOLR2 expression in Kupffer cells (CD11b^int^F4/80^+^TIM-4^+^) and their impaired phagocytosis function by RRV infection was also restored by CF treatment (Figure 4L). We also found histological evidence of improvement in CF-treated than non-treated BA mice as indicated by the reduction in steatosis, septal fibrosis, necrosis foci, lymphocyte accumulation around the portal tracts and epithelial vacuolation/swelling (Figure 4M).

### Folic acid suppresses IFN-hepcidin expression and restores iron and redox balance

*FOLR2* encodes a receptor that facilitates unidirectional transport of folate and reduced folic acid derivatives and mediates delivery of 5-methyltetrahydrofolate into the interior of cells. Given that *FOLR2* expression was significantly decreased in the hepatic and intestinal myeloid subsets in BA compared with control-CC subjects (Figure 4E and 4F), we wanted to determine whether FOLR2 may regulate IFN-I signaling and hepcidin expression (Figure 5A). First, we used M-CSF differentiated bone marrow derived macrophages (BMDM). We stimulated BMDM at day 5 with Poly(dA:dT) to mimic viral infection ^39^, and treated them with or without folic acid for 18 hours (Figure 5B). We found that FOLR2 was expressed in a time-dependent manner in M-CSF differentiated macrophages (Figure 5C). Poly(dA:dT) treatment promoted IFN-I and downstream genes (*Ifna1*, *Ifnb*, *Irf1* and *Ifit2*) (Figure 5D), as well as *Hamp* and *Trem2* expression (Figure 5E). In contrast, *Folr2* expression was decreased by Poly(dA:dT) treatment, but this trend was reversed following folic acid supplementation (Figure 5E). These data suggest that viral infection may promote hepcidin-expressing TREM2^+^Mac differentiation. In contrast, folic acid supplementation favours FOLR2^+^Mac differentiation.

**Figure 5.**
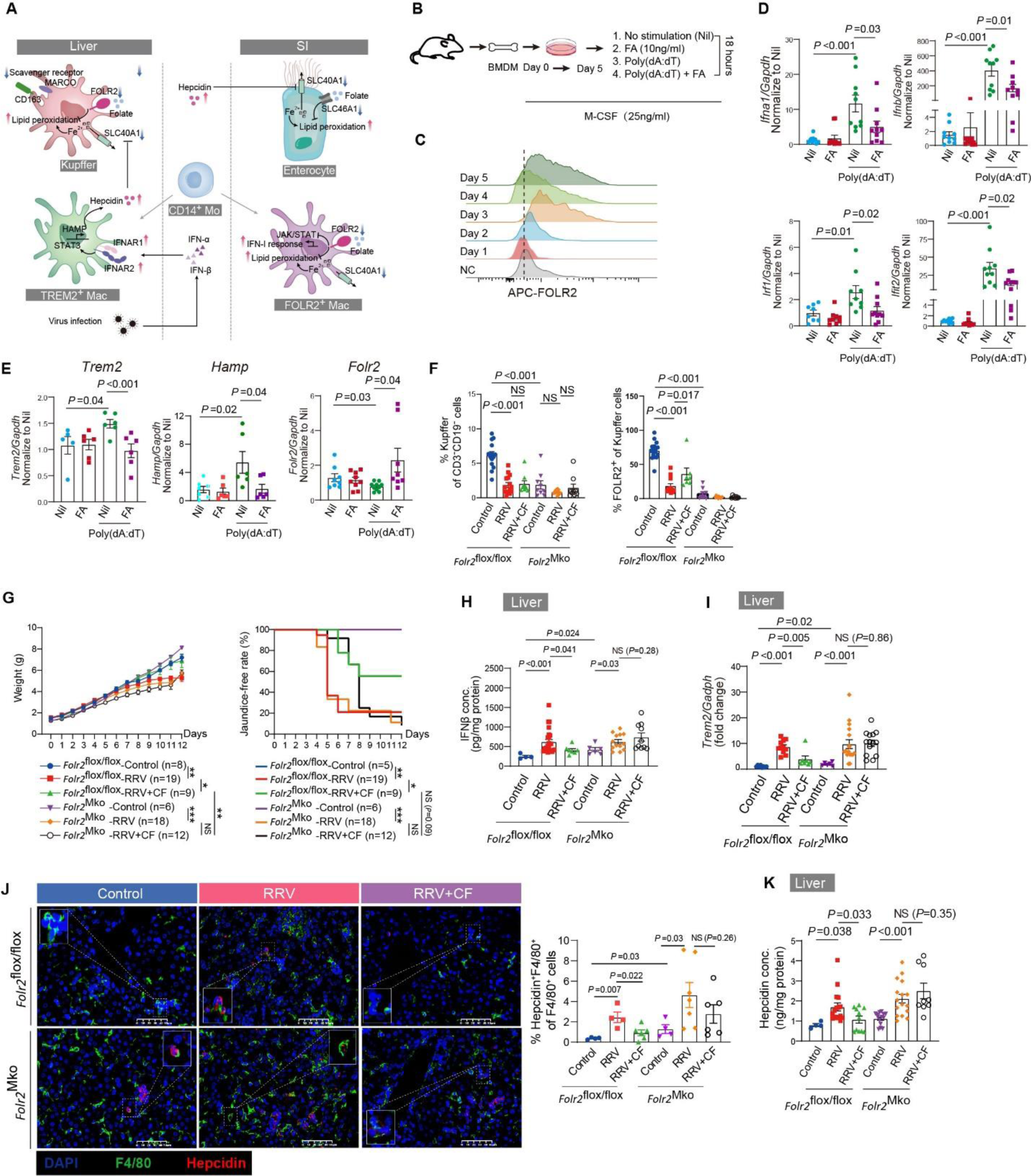
Folic acid suppresses IFN-hepcidin expression and restores iron and redox balance. (**A)** Schematic diagram depicting interactions of hepatic and intestinal macrophages with intestinal epithelial cells. (**B**) Schematic for *in vitro* culture and stimulation of bone marrow derived macrophages (BMDM). (**C**) FOLR2 expression was measured by flow cytometry during bone marrow differentiation. NC represents for negative control without APC-FOLR2 staining. (**D** and **E**) Relative expression of genes were examined by qPCR in BMDM transfected with Poly (dA: dT) with or without folic acid (FA) treatment. Data were normalized to the relative expression of indicated genes in Nil (unstimulated) conditions. (**F**) The frequencies of Kupffer cells and FOLR2^+^ Kupffer cells were measured by flow cytometry, gating strategies were shown in **Supplementary Figure. 7B**. (**G**) Line charts displaying body weight changes and jaundice occurrence rates. The statistics for body weight change was performed using Two-Way ANOVA, for jaundice-free survival rate and survival probability were performed using Log-Rank test; *p < 0.05, **p < 0.01, ***p < 0.001. (**H**) Dot plots showing concentrations of murine IFN-β in liver homogenates, shown as per mg protein. (**I**) Relative expression of *Trem2* in liver were examined by qPCR. (**J**) Representative immunofluorescent images showing co-staining of F4/80 (green) and hepcidin (red) in liver biopsies. Scale bar, 100μm. Dot plots showing percentage of F4/80^+^hepcidin^+^ cells of total F4/80^+^ cells. (**K**) Dot plots showing hepatic hepcidin concentration. For all data, each point represents an individual patient/mouse, and the line represents the mean ± SEM. *P* values were calculated by two-tailed Student’s t test unless otherwise indicated.

To further reveal the contribution of folate on macrophage function *in vivo*, we generated myeloid-specific *Folr2* knockout mice (*Folr2^MKO^*) on a BALB/c background (Figure S7A). Conditional *Folr2* knock-out was confirmed by PCR genotyping and flow cytometry as shown in Figure S7B and S7C. The expression of FOLR2 was markedly reduced in Kupffer cells (CD11b^int^F4/80^+^), monocytes (CD11b^+^Ly6C^hi^) and neutrophils (CD11b^+^Ly6G^hi^) in the livers of *Folr2^MKO^* mice compared with *Folr2^flox/flox^*mice (Figure 5F; Supplementary Figure 7C-D). We observed that the frequencies of Kupffer cells were already significantly decreased in the livers of *Folr2^MKO^* mice compared with *Folr2^flox/flox^* mice under PBS-treated conditions, which suggest that FOLR2 is required for Kupffer cell development or survival in the neonatal mice. In *Folr2^flox/flox^* control littermates, we found that RRV infection decreased the expression of FOLR2 in Kupffer cells (Figure 5F) and increased monocyte infiltration (Supplementary Figure 6C-D). Calcium folinate (CF) treatment partially restored FOLR2 expression in Kupffer cells in the *Folr2^flox/flox^* control littermates (Figure 5F; Supplementary Figure 7C).

We next investigated whether myeloid-expressed FOLR2 mediated some of the therapeutic effects of CF. We showed that CF treatment significantly ameliorated jaundice incidence, weight loss (Figure 5G) and liver and small intestine pathology (Supplementary Figure 8A-B) in RRV-infected *Folr2^flox/flox^*littermate mice. However, these therapeutic effects of CF were lost in the RRV-infected *Folr2^Mko^* mice (Figure 5G). In BMDM obtained from *Folr2^Mko^* mice, folic acid treatment did not inhibit the expression IFN-I and IFN-I-stimulated genes and *Hamp* (Supplementary Figure 8C). Consistently, the therapeutic effects of CF were associated with decreased IFN-I (Figure 5H), decreased differentiation of hepcidin-expressing TREM2^+^Mac cells (Figure 5I and 5J), and decreased hepcidin expression (Figure 5K) in the liver of *Folr2^flox/flox^* littermate compared with *Folr2^Mko^* mice. Consequently, CF treatment increased SLC40A1 expression in the small intestine (Supplementary Figure 8D), increased iron excretion (Supplementary Figure 8E) and decreased iron accumulation (Supplementary Figure 8F) and ROS production (Supplementary Figure 8G) in the liver in *Folr2^flox/flox^* littermates compared with *Folr2^Mko^*mice. Collectively, these data suggest that CF decreases jaundice incidence by suppressing IFN-I and hepcidin expression in myeloid cells via interaction with FOLR2.

### Folic acid supplementation reduces jaundice incidence, cholangitis incidence and liver transplantation rate in infants with BA

Given the above success in RRV-induced BA mice, we investigated whether folic acid supplementation may improve the prognosis of infants with BA in an open label clinical trial. We administered folic acid instead of its active metabolite (folinate) because 1) only folic acid is available for oral administration in our medical center, and 2) adequate folic acid supplementation has been shown to bypass SLC46A1 in patients with hereditary folate malabsorption ^40^. Nineteen subjects with BA were given oral folic acid at 0.4mg/day for 6 months (Figure 6A). A group of infants with BA (n=38) matched for age (at Kasai’s procedure), gender, liver enzyme, bile acid and bilirubin status, and operation surgeons were selected to compare folic acid treatment efficacy (Figure 6B). Significant decreases in total and direct bilirubin concentrations were observed for folic acid-treated patients starting from 3 months follow-up (Figure 6D and 6E). At the 6-month follow-up, the overall incidence of bacterial cholangitis decreased from 74% (28/38) in the untreated to 21% (4/19) in the folic acid treated patients (Figure 6C; Supplementary Figure 9A). At 2-years follow-up, the rates of liver transplantation decreased from 41.1% (14/34, 4 participants lost contact during follow-up) in the untreated to 11.1% (2/18, 1 participants lost contact during follow-up) in the folic acid treated patients (Figure 6F).

**Figure 6.**
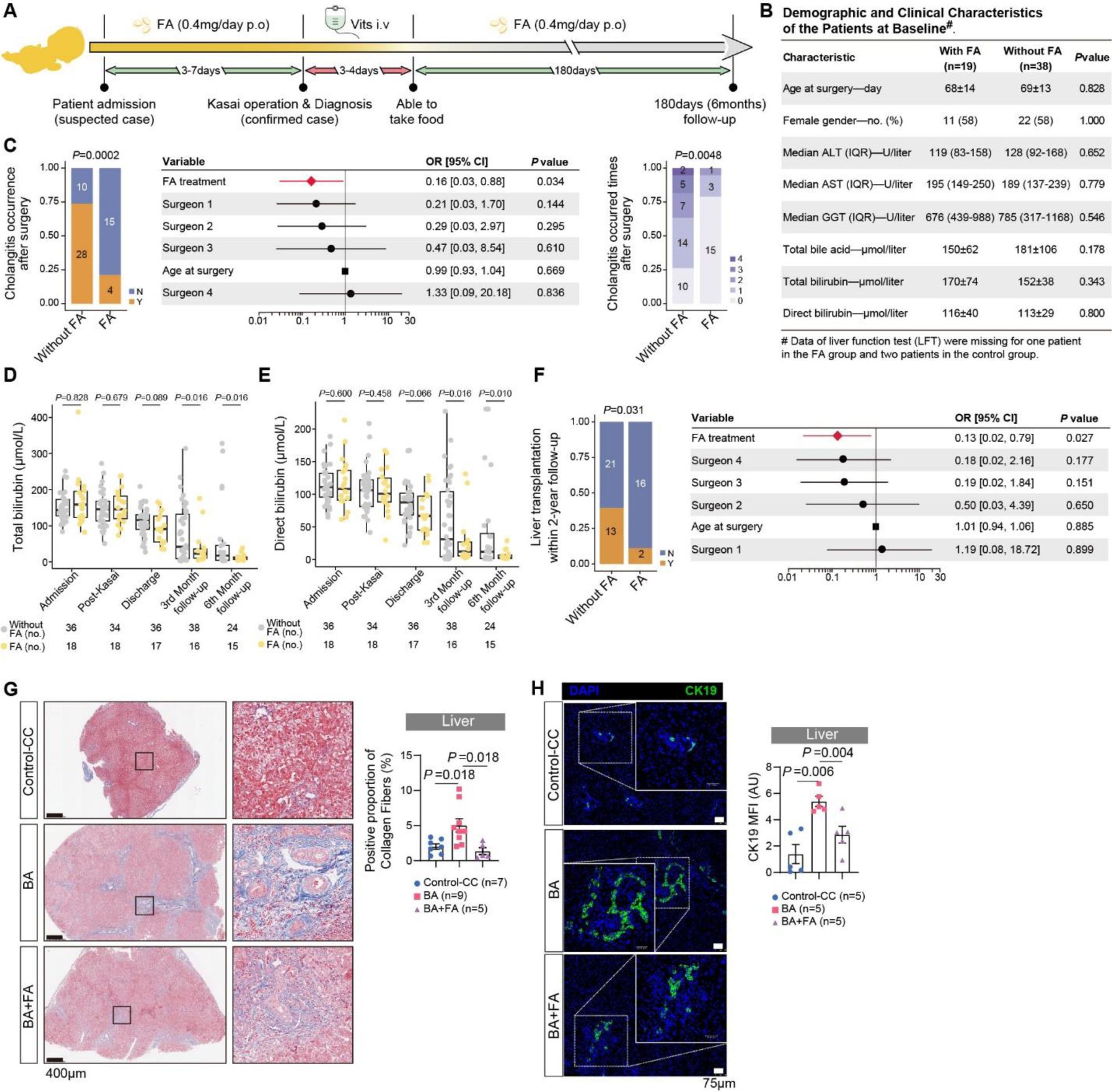
Folic Acid Supplementation Improves Jaundice Clearance rate, Reduces Cholangitis Incidence, and Improves Self-liver Survival in Infants with BA. (**A**) Schematic diagram depicting folic acid (FA) administration protocol in infant with BA. (**B**) Table summarizing demographic and clinical characteristics of the FA-treated and un-treated patients at baseline. Data are presented as mean ± SD or mean (range). (**C**) Stacked bar plots at left panel comparing the proportions of postoperative cholangitis occurrence within the 6-month follow-up for patients with or without folic acid treatment. *P* value was determined by Fisher’s exact test. Table in the middle summarizes effects of folic acid treatment, different surgeons, gender and age at surgery on the result of postoperative cholangitis occurrence. *P* value for each factor was determined by Likelihood ratio test, and the significant factor was labeled as red. Stacked bar plots at right panel showing occurrence times (shown as different colors) of postoperative cholangitis within 6-month follow-up of the patients with or without FA treatment. *P* value was determined by Chi-squared test. (**D** and **E**) Total bilirubin (TBIL) and direct bilirubin (DBIL) levels in patients with or without FA treatment at different follow-up time points were recorded. *P* values were determined by Mann-Whitney U test. Bar plots showing the relative changes of TBIL and DBIL of patients at 3-month FA treatment in comparison with the baseline. (**F**) Proportions of liver transplantation within the 2-year follow-up for patients with or without FA treatment were shown in stacked bar plots. *P* value was determined by Fisher’s exact test. Table showing effects of FA treatment, different surgeons, gender and age at surgery on the result of liver transplantation of patients with BA. (**G** and **H**) Representative hematoxylin-eosin (H&E) and Masson’s trichrome stained sections for liver biopsies. Quantification of Masson’s trichrome staining using Image-J software was shown as percentage of collagen staining area per mm^2^ visual field. (**I**) Immunofluorescent staining of CK19 in liver biopsies. Scale bars, 75μm. For all data, each point represents an individual patient, and the line represents the mean ±SEM. *P* values were calculated by two-tailed Student’s t test unless otherwise indicated.

We then performed H&E and Masson’s trichrome staining for liver biopsies and found that folic acid treatment decreased infiltration of immune cells at the portal area (Figure 6G). Masson’s trichrome staining intensity was dramatically decreased, which suggested that folic acid is effective in prevention of liver cirrhosis (Figure 6H). Decreased anti-CK19 immunohistochemical staining indicated reduction of intrahepatic bile duct hyperproliferation (Figure 6I). These changes were consistent with significantly decreased liver transplantation rate 2 years post-Kasai’s operation in BA treated with folic acid (Figure 6F).

To confirm the underlying mechanisms we described above, we also generated an additional scRNA transcriptome dataset using the BD Rhapsody platform (Supplementary Figure 9B) in a separate control-CC (n=4), BA with (n=5) or without (n=8) folic acid treatment. Among the 12 epithelial cell subsets (Figure 7A), genes that regulate folic acid absorption (*SLC46A1*, *FOLH1*), iron export (*FTH1*, *NCOA4*, *SLC40A1*) and suppression of lipid peroxidation (*NFE2L2*/*NRF2*, *SELENOP*, *GPX4*) were significantly upregulated in the intestinal epithelial cells in folic acid treated compared with untreated BA patients (Figure 7B; Supplementary Figure 9C). In contrast, the response to IFN-I (*STAT1*, *IFITM2*, *IFITM3*) was significantly decreased in intestinal epithelial cells (Figure 7B; Supplementary Figure 9C). Suppression of IFN-I signaling was also observed in myeloid cells from both the small intestine and liver (Figure 7C-D). TREM2^+^Mac from the folic acid treated patients displayed decreased expression of *HAMP* and *TREM2* (Figure 7D; Supplementary Figure 9D), whereas Kupffer cells showed increased scavenger function (*SCARF1*) and erythrophagocytosis (*CD163*, *MFGE8*, *CLEC4F*, *CYP3A4*) (Supplementary Figure 9E). We confirmed these findings using tissue biopsies obtained from patients receiving Kasai’s procedure, and showed that folic acid treatment significantly upregulated the expression of NRF2 and GPX4 (Supplementary Figure 9F), decreased production of IFN-I cytokines (IFN-α, IFN-β) and hepcidin (Figure 7E).

**Figure 7.**
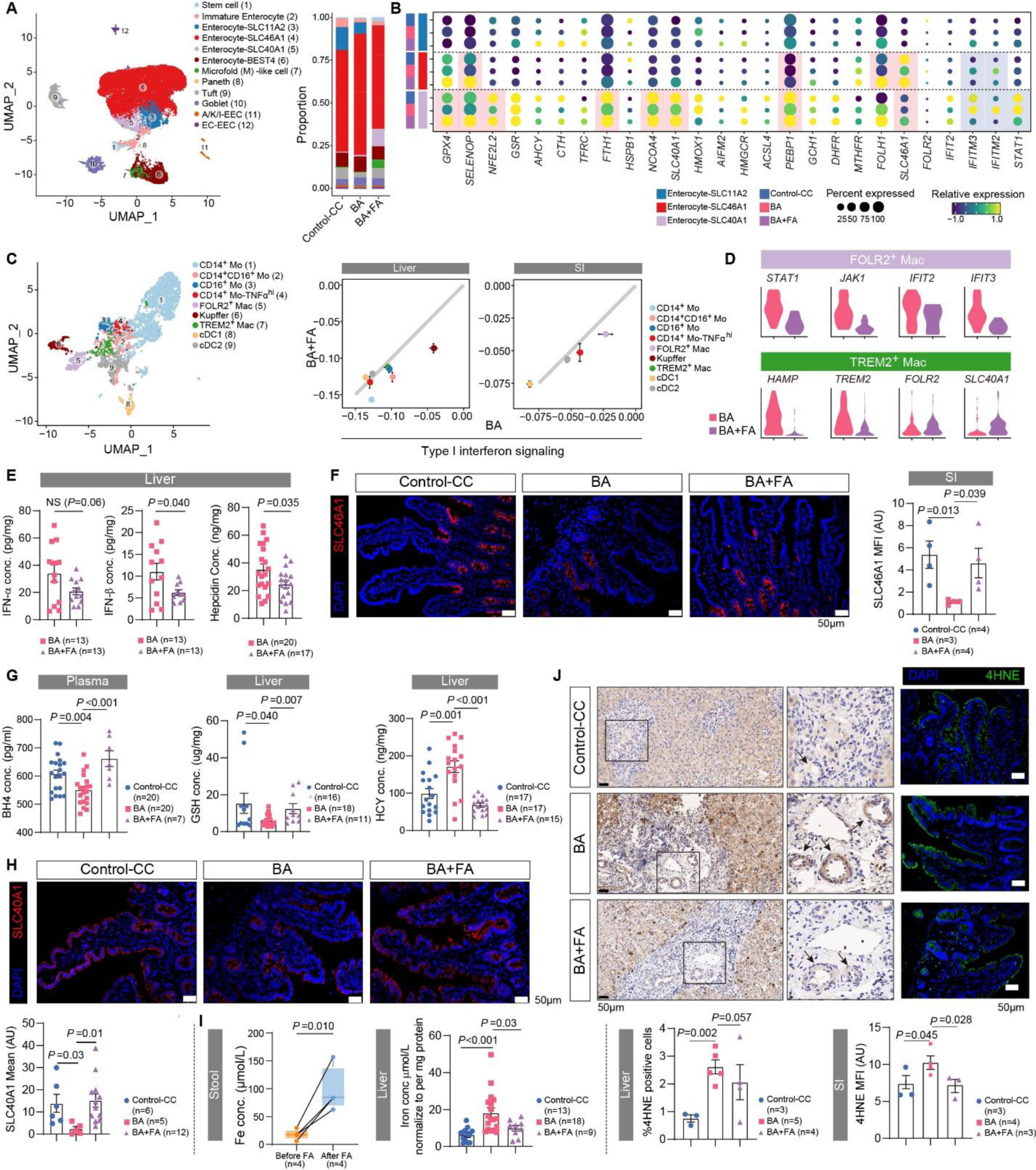
FA Supplementation Restores IFN-I-Iron and Redox balances in Infants with BA. (**A)** UMAP plots displaying distributions of 12 subsets of enterocytes. (**B**) Gene expression across Enterocyte-SLC11A2, Enterocytes-SLC46A1 and Enterocytes-SLC40A1 was compared among patients with CC, BA and BA with FA treatment. (**C**) UMAP plots displaying distributions of 9 clusters of myeloid. Scatter plots comparing the signature scores of IFN-I signaling for each subset of myeloid cells in BA subjects (x-axis) and BA with FA treatment (y-axis). (**D**) Violin plots showing relative gene expression in intestinal FOLR2^+^Mac and TREM2^+^Mac between BA with and without FA treatment. (**E**) Dot plots showing IFN-α, IFN-β and hepcidin in liver homogenates. (**F**) Immunofluorescent staining of SLC46A1 in small intestine. Scale bars, 50μm. (**G**) Dot plots showing BH4 in plasma, GSH and HCY in liver homogenates. (**H**) Immunofluorescent staining of SLC40A1 in small intestine. Scale bars, 50μm. (**I**) Box plots comparing preoperative changes of iron concentration in stool samples from 4 patients with BA before and after FA treatment, shown as per mg weight. Dot plots showing concentrations of total iron in liver homogenates. (**J**) Representative immunohistochemistry images showing 4-HNE expression in liver biopsies. Quantification was percentage of 4-HNE^+^ cells per mm^2^ visual field. Scale bar, 50μm. Right panel: representative immunofluorescent images showing 4-HNE staining in the small intestine biopsies. Scale bar, 50μm. *P* values were calculated by two-tailed Student’s t test for all experiments unless otherwise indicated.

Folic acid treatment also restored SLC46A1 expression in intestinal epithelial cells (Figure 7F). Such treatment also significantly increased the concentrations of circulating tetrahydrobiopterin (BH4) and hepatic GSH, but decreased the concentrations of hepatic homocysteine (Figure 7G), confirming that folic acid supplementation restored folic acid absorption and redox balance ^41, 42^. The corrections in systemic iron accumulation and luminal iron deficiency were evidenced by increased SLC40A1 expression in intestinal epithelial cells (Figure 7H), increased stool iron levels and decreased systematic iron accumulation in the preoperative folic acid treated patients (Figure 7I, Supplementary Figure 9G). Consequently, lipid peroxidation (measured by 4-HNE expression) was significantly decreased in the small intestine and liver in folic acid treated BA patients (Figure 7J). At the adaptive immunity level, folic acid treatment reduced IgG-Ro/SSA autoantibody production by hepatic B cells, and increased IgM/IgG4 ratio (Supplementary Figure 9H), which we have previously shown to correlate with prolonged self-liver survival in infants with BA ^21^.

#### Genes that Regulate IFN-I signaling and Folic Acid Metabolism are Associated with BA Risk

Finally, we wanted to determine whether genetic predisposition may promote BA risk in infants. We performed a genome-wide association study for infants with BA (n=281) and control (n=3453) subjects. Gene-based assays were used to identify candidate risk variants ^43^. We found that genes (Supplementary table 2 and table 3) that regulate intestinal epithelial integrity (*PIGR*, *WFDC2*), IFN-I signaling (*ISG20*, *PLD3*, *SOCS3*, *STAT1*, *IFNAR2*, *OAS1*), iron and folate metabolism (*AHCY*, *NFE2L2*, *GPX4*, *COQ4*, *HAMP*, *ISCU*, *SHMT2*) were differentially enriched in infants with BA compared with non-BA control subjects (Supplementary Figure 10).

## Discussion

Biliary atresia (BA) is a multiple factorial disease with obliterative extrahepatic cholangiopathy as the common endpoint. Viral infection has been considered an important trigger for disease development but the molecular mechanism leading to disease development remained elusive. In this study, we report that persistent IFN-I activation, high likely derived from a perinatal viral infection, promotes systemic iron accumulation and intestinal lumen iron deficiency by hepcidin-induced SLC40A1 degradation. Under oxidative stress conditions, iron accumulation promotes lipid peroxidation and ferroptosis. Therefore, blocking persistent IFN-I -JAK-STAT signaling, or iron chelation, reduced BA incidences in the RRV-induced BA model. In addition, we show that viral infection suppresses folic acid absorption and metabolism in infants (in BA and HCMV^+^NH) and neonatal mice, which in turn promotes persistent IFN-I activation. Our collective findings suggest that neonatal viral infection may induce persistent IFN-I activation and folic acid deficiency in at-risk individuals, which subsequently leads to systemic iron accumulation, oxidative stress, lipid peroxidation and ferroptosis in infants with BA. This is shown schematically in Supplementary Figure 11.

Folic acid is an essential nutrient that is critical for early life development. It provides one carbon unit for the biosynthesis of DNA, proteins, lipids, and epigenetic modification of the genome ^44^. It also plays essential roles in redox maintenance by promoting synthesis of glutathione (GSH) and tetrahydrobiopterin (BH4) (via DHFR) ^45, 46^. Prenatal folate supplementation prevents congenital neurological and heart defects, as well as low birthweight ^47–49^. We show that post-natal viral infection leads to folic acid malabsorption and demonstrate that folic acid supplementation ameliorates virally associated immune pathology. In an open label clinical trial, folic acid treatment markedly increased jaundice clearance rate, decreased bacterial cholangitis incidence, and most strikingly, significantly improved self-liver survival in infants with BA. Viral infection is associated with the development of a spectrum of autoimmune diseases in childhood ^50^, including HCMV^+^ NH. Given that IFN-I, iron and folic acid were similarly altered in infants with HCMV^+^NH compared with infants with BA, it would be interesting to determine whether folic acid supplementation may provide therapeutic benefits in virally induced hepatic and intestinal diseases in infants and children.

Our data suggest that at least three factors have contributed to folic acid deficiency in infants with BA. First, infection and treatment related microbial dysbiosis decrease intestinal folate availability. Second, viral infection induces inefficient folic acid absorption due to deceased SLC46A1 and FOLR2 expression by the small intestinal epithelial cells and tissue macrophages. Third, decreased DHFR expression impairs folic acid metabolism. By generating a myeloid-specific *Folr2* knockout mice, we demonstrated that folic acid played a critical role in Kupffer and FOLR2^+^ macrophage differentiation. Specifically, we showed that Kupffer cells were decreased in numbers even under uninfected conditions in the *Folr2^Mko^* mice. Previously, we have shown that the scavenging function of Kupffer cells was impaired in infants with BA compared with control-CC subjects ^21^. In this study, we showed that folic acid supplementation in infants with BA restored erythrophagocytosis function of Kupffer cells. In addition, our data suggest that folic acid suppresses the differentiation of hepcidin-expressing TREM2^+^mac from bone marrow derived monocytes upon viral infection. The treatment efficacy of calcium folinate found in RRV-infected wildtype mice was diminished in RRV-infected *Folr2^Mko^* mice, which presented with increased IFN-I expression and iron accumulation. Further studies are still required to determine the molecular aspects on how folic acid regulates tissue and circulating macrophage differentiation and function.

Folic acid treatment exhibits several important therapeutic benefits for infants with BA. First, folic acid suppresses persistent IFN-I signaling and reduces hepcidin expression by TREM2^+^Mac cells. This restores systemic iron balance by increasing SLC40A1-mediated luminal iron excretion. Second, folic acid increases anti-oxidative function by increasing GPX4 and SELENOP expression, increasing plasma BH4, hepatic GSH and decreasing hepatic homocysteine concentrations. These changes reduce lipid peroxidation and ferroptosis of the small intestine and liver in folic acid treated patients. Third, folic acid decreases autoantibody production by hepatic B cells. Collectively, folic acid treatment achieved remarkable clinical outcomes for BA as exemplified by expedited bilirubin clearance by 3 months, dramatically reduced cholangitis incidence by 6 months, and strikingly improved self-liver survival rate by 1.5 years post Kasai’s operation. Placebo controlled double blinded clinical trials should be followed to confirm the therapeutic benefits of folic acid to biliary atresia. Preventive clinical trials may also be considered in at risk neonates for reducing virally-induced BA development in infants.

## Data Availability

All data produced in the present work are contained in the manuscript.

https://ngdc.cncb.ac.cn/gsa-human

## Grant support

The National Natural Science Foundation of China (92042303 (Y.Z.), 82125015 (Y.Z.), 82201892 (Y.X.), 82201891 (X.C.), 82003431 (X.T.), 82101808 (Z.L.), 82241220 (Zhi Yao), 82001589 (Zhenhua Luo), 92168108 (Zhenhua Luo)), Basic and Applied Basic Research Foundation of Guangdong Province 2021B1515230003 (X.G.), Basic and Applied Basic Research Foundation of Guangdong Province 202102080511(Q.H.), Natural Science Fund of Guangdong Province (202102020829 (X.G.), 2022A1515012558 (X.G.)), Research Fund of Guangzhou Women and Children’s Medical Center (2020LH-4-005 (J.L.)) funded this study.

## Disclosures

Guangzhou Women and Children’s Medical Center has filed patent applications (The application of folic acid in the diagnosis, prevention, and treatment of hereditary, infectious or allergic diseases, 202211377355.3, 11/4/2022). Y.Z., Z.W., Y.X., Z.C., X.C., and R.F. are named inventors of the patent. The authors do not have any paid or unpaid consulting to declare.

## Author contributions

Conceptualization: Y.Z. Methodology: Z.W., Y.N., J.L., Q.L., C.C., Q.H., Z.L., Yi Xu, and K.L. (recruited, provided patient care and collected follow-up data), Y.X., R.F., W.L., X.L., and Yunnan Xiao (laboratory experiments), Y.H. (clinical survey), Z.L. and Zhenhua Luo (mouse model construction), Z.C., X.C., and Yanfang Zhang (bioinformatic analysis), X.L., H.R., X.T. and Q.Y. (collection of patient specimens), K.Z. (RNA-seq library construction for 10x Genomics single cell 5’ and V(D)J sequencing and for BD Rhapsody™ Single-Cell Analysis System), Z.Z., L.S., X.Z. and Y.Z. (GWAS analysis). Writing-review and editing: Y.Z., Y.X., A.M.L., Z.C., X.C., R.F., J.L., H. X., W. Z. and Z.Y.. All authors read and approved the final manuscript.

## Data and materials availability

The raw sequence data reported in this paper have been deposited in the Genome Sequence Archive (Genomics, Proteomics & Bioinformatics 2021) in National Genomics Data Center (Nucleic Acids Res 2022), China National Center for Bioinformation / Beijing Institute of Genomics, Chinese Academy of Sciences (GSA-Human: HRA005080) that are publicly accessible at https://ngdc.cncb.ac.cn/gsa-human upon approval. All data associated with this study has been presented in the paper and the Supplementary Materials.

## Notes

### Competing Interest Statement

The authors have declared no competing interest.

### Clinical Trial

Chinese clinical trial, ChiCTR2100050992

### Funding Statement

This study was funded by The National Natural Science Foundation of China,Basic and Applied Basic Research
Foundation of Guangdong Province, Basic and Applied Basic Research Foundation of Guangdong Province, Natural Science Fund of Guangdong Province, Research Fund of Guangzhou Women and Children Medical Center.

### Author Declarations

Medical Ethics Committee of Guangzhou Women and Children's Medical Center gave ethical approval for this work.

