## Supplementary Figures and legends for "Folic acid prevents interferon-induced iron accumulation and ferroptosis and improves liver health in children with biliary atresia"

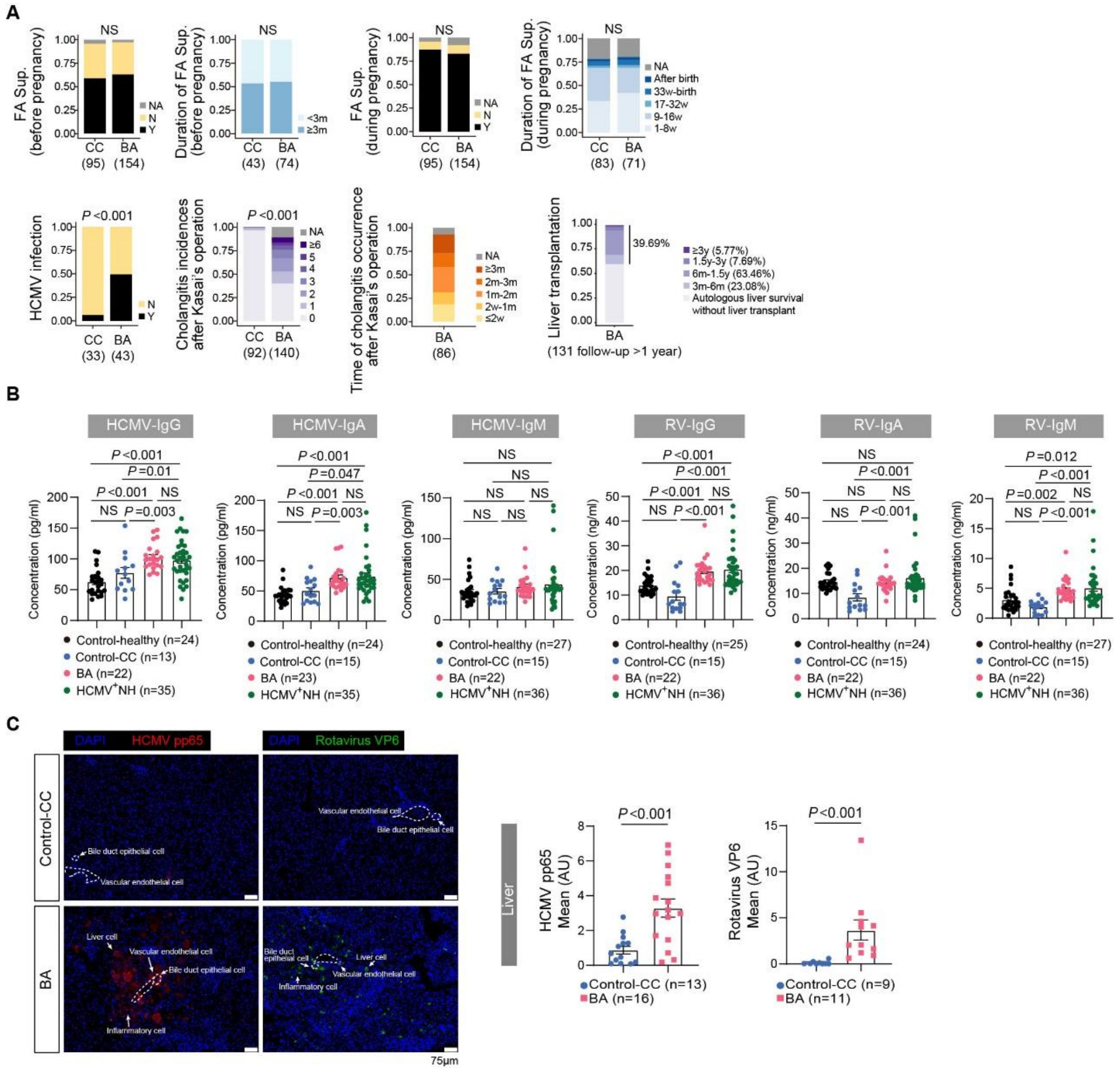


**Supplementary Figure 1. Presence or history of viral infection in infants with BA.** (**A**) Stacked bar plots showing survey results for control-CC subjects and infants with BA. Representations of colored bars are indicated. P values were determined by Chi-Squared test. FA, folic acid; HCMV, human cytomegalovirus. (**B**) Serum levels of IgM, IgG and IgA antibodies specific for human rotavirus (RV) and HCMV in age matched control-healthy, control-CC, BA and HCMV$^{+}$ NH were measured by ELISA. Each point represents an individual patient, and the line represents the mean ± SEM. (**C**) Immunofluorescence analysis for HCMV pp65 (red) and RV VP6 (green) in liver biopsies from control-CC and infants with BA (n=16). Quantitation (mean ± SEM) and representative pictures are shown. Each point represents an individual patient. Scale bars, 75 μm. *P* values were calculated by two-tailed Student's t test for all experiments.

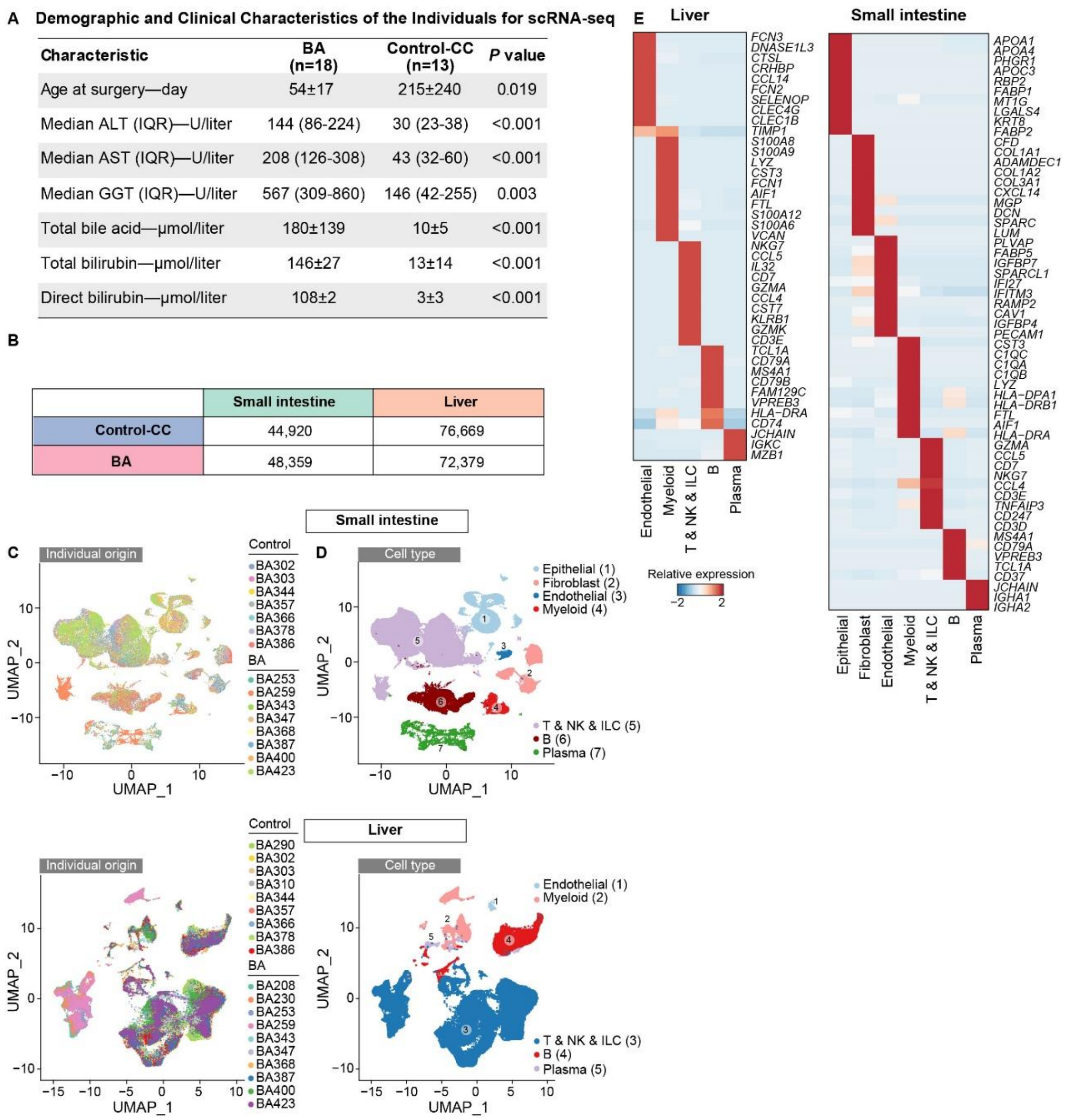

**A Demographic and Clinical Characteristics of the Individuals for scRNA-seq**

| Characteristic | BA (n=18) | Control-CC (n=13) | *P* value |
|---|---|---|---|
| Age at surgery—day | 54±17 | 215±240 | 0.019 |
| Median ALT (IQR)—U/liter | 144 (86-224) | 30 (23-38) | <0.001 |
| Median AST (IQR)—U/liter | 208 (126-308) | 43 (32-60) | <0.001 |
| Median GGT (IQR)—U/liter | 567 (309-860) | 146 (42-255) | 0.003 |
| Total bile acid—μmol/liter | 180±139 | 10±5 | <0.001 |
| Total bilirubin—μmol/liter | 146±27 | 13±14 | <0.001 |
| Direct bilirubin—μmol/liter | 108±2 | 3±3 | <0.001 |

**B**

| | Small intestine | Liver |
|---|---|---|
| Control-CC | 44,920 | 76,669 |
| BA | 48,359 | 72,379 |



**Supplementary Figure 2. Single-cell RNA profiling of small intestine and liver biopsies for control subjects and infants with BA.** (**A**) Table summarizing demographic and clinical characteristics of individuals for scRNA-seq. Data are presented as mean±SD or mean (range). (**B**) Table summarizing cell numbers generated by 10X scRNA-seq for small intestine and liver from control-CC and patients with BA. (**C** and **D**) UMAP plots displaying distributions of major cell types and sample origins at single cell resolution. (**E**) Marker gene expression was shown for 7 major cell clusters from the small intestine and 5 major cell clusters from the liver.

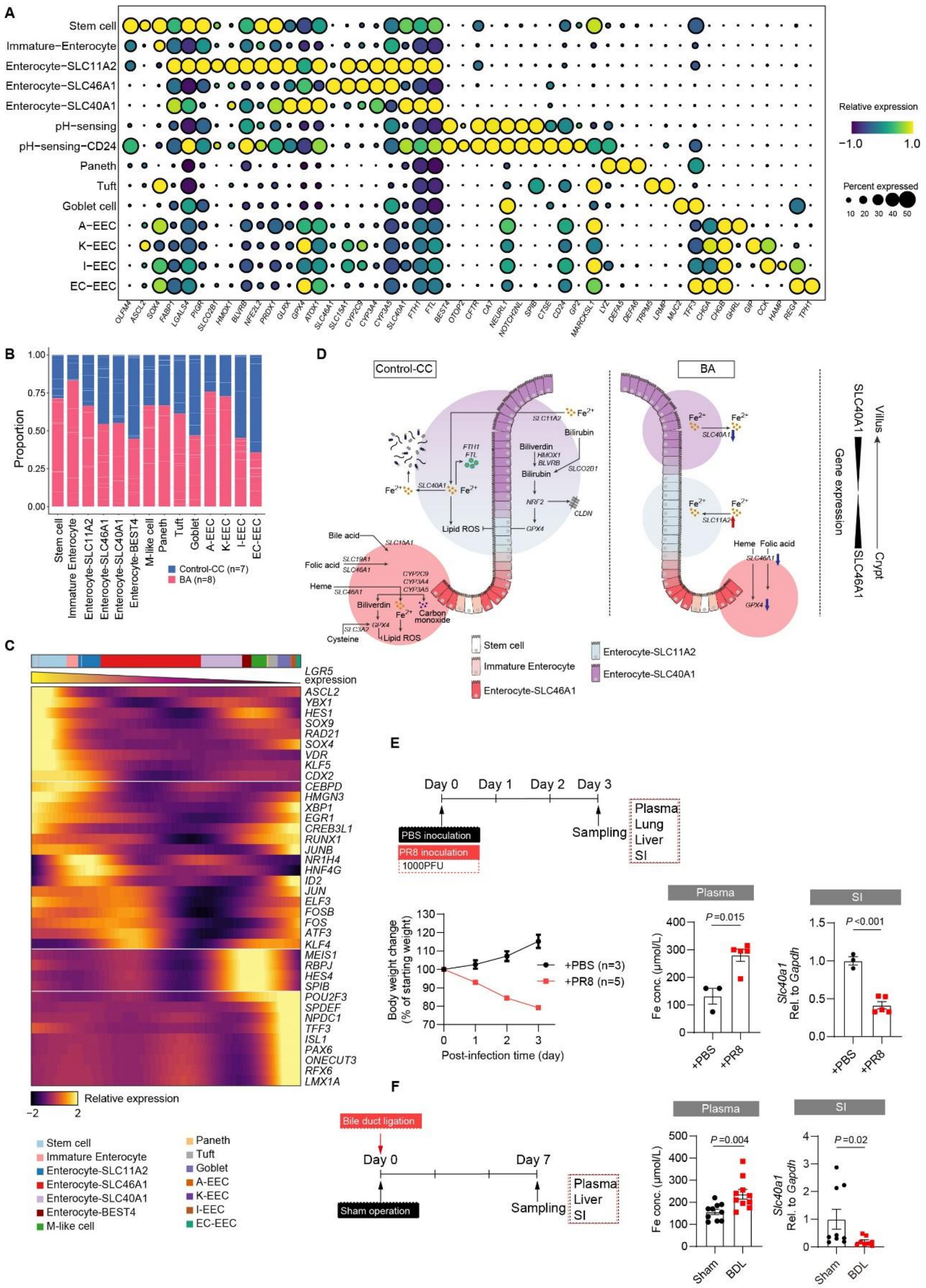


**Supplementary Figure 3. Absorption, storage, and excretion of iron in control subjects and infants with BA.** (**A**) Marker gene expression was shown across 14 cell subsets of the small intestinal epithelium. (**B**) Bar plots showing cell subset distributions across samples within different groups. Blocks represent individual samples. (**C**) Heatmap displaying smoothing spline curves of differentially expressed transcription factors across different small intestinal enterocyte subsets. The spline curves of genes were fitted to LGR5 expression. (**D**) Schematic diagram depicting distribution of SLC40A1, SLC11A2 and SLC46A1 expressing enterocytes and metabolic pathways of heme, iron and folic acid in control-CC subjects and infants with BA. (**E**) Schematic for PR8 virus inoculation in 14-day-age mice of BALB/C background. Body weight were monitored daily after infection, and mice were euthanized when weight loss reached 20%. Plasma iron content was measured. Gene expression of *Slc40a1* in small intestine biopsies was estimated by qPCR. Line represents the mean ± SEM. *P* values were calculated by two-tailed Student's t test. (**F**) Schematic for mouse model of bile duct ligation (BDL). Mice in sham group underwent common bile duct dissociation but no ligation. Plasma iron content was measured. Gene expression of *Slc40a1* in small intestine biopsies was estimated by qPCR. Line represents the mean ± SEM. *P* values were calculated by two-tailed Student's t test.

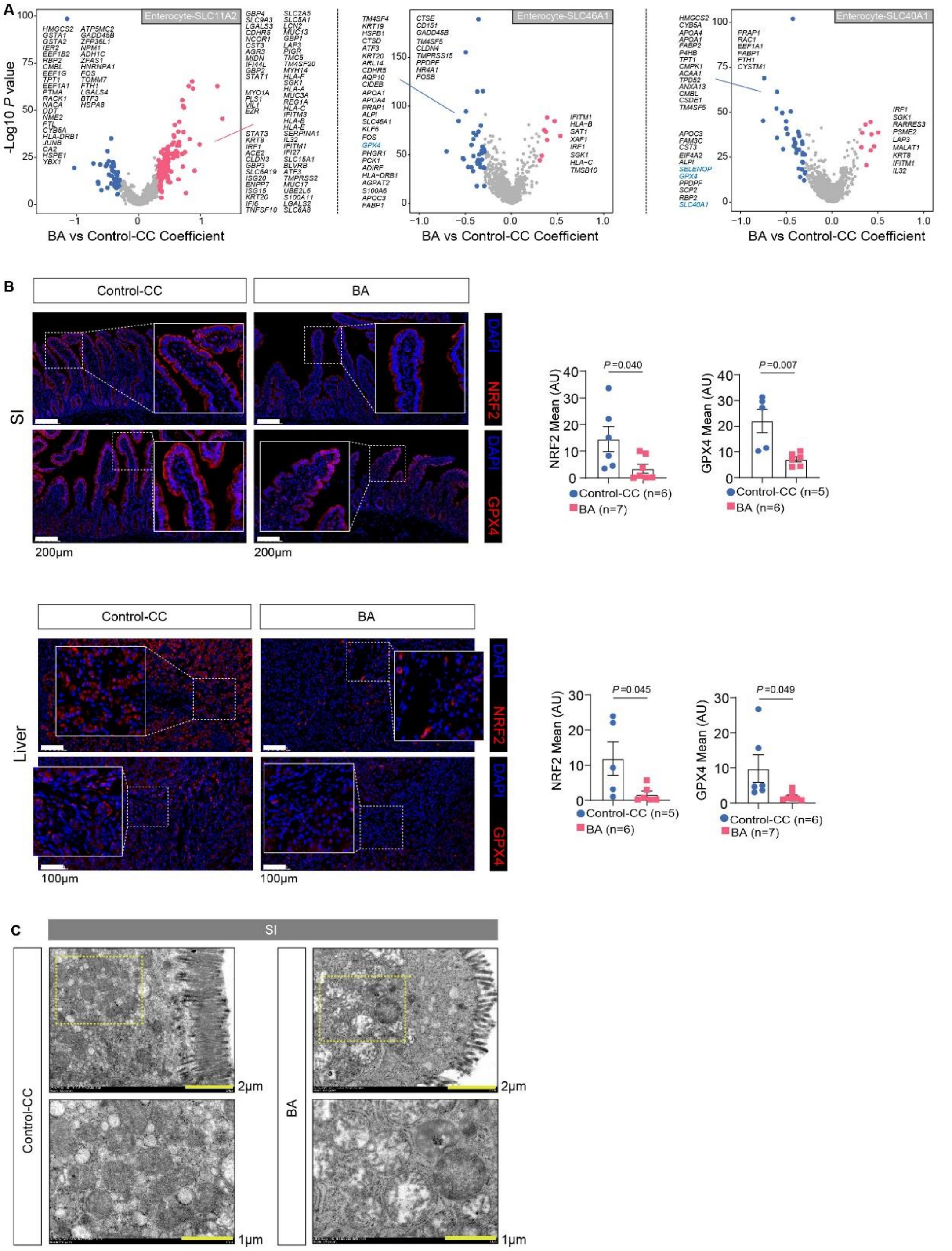


**Supplementary Figure 4. Elevated IFN-I and decreased anti-oxidative responses in infants with BA.** (**A**) Volcano plots showing differentially expressed genes of enterocyte-SLC11A2, enterocyte-SLC40A1 and enterocyte-SLC46A1 in comparison to their counterparts from control-CC subjects. Red and blue dots denote the up-regulated and down-regulated genes passing the thresholds of P value <=0.05 and |Coefficient| >=0.25, respectively. (**B**) Representative immunofluorescent images showing GPX4 and NRF2 staining in liver and small intestine biopsies from BA and control-CC subjects. Scale bar, 100μm for liver specimens and 200μm for small intestine specimens. Measurement of mean fluorescence intensity (MFI) for NRF2 and GPX4 and the statistical unit was shown as Arbitrary Unit (AU). *P* values were calculated by two-tailed Student's t test. (**C**) Representative transmission electron micrographs of ultrathin intestinal sections comparing BA (n=4) and control subjects (n=4), Scale bar, 1μm.

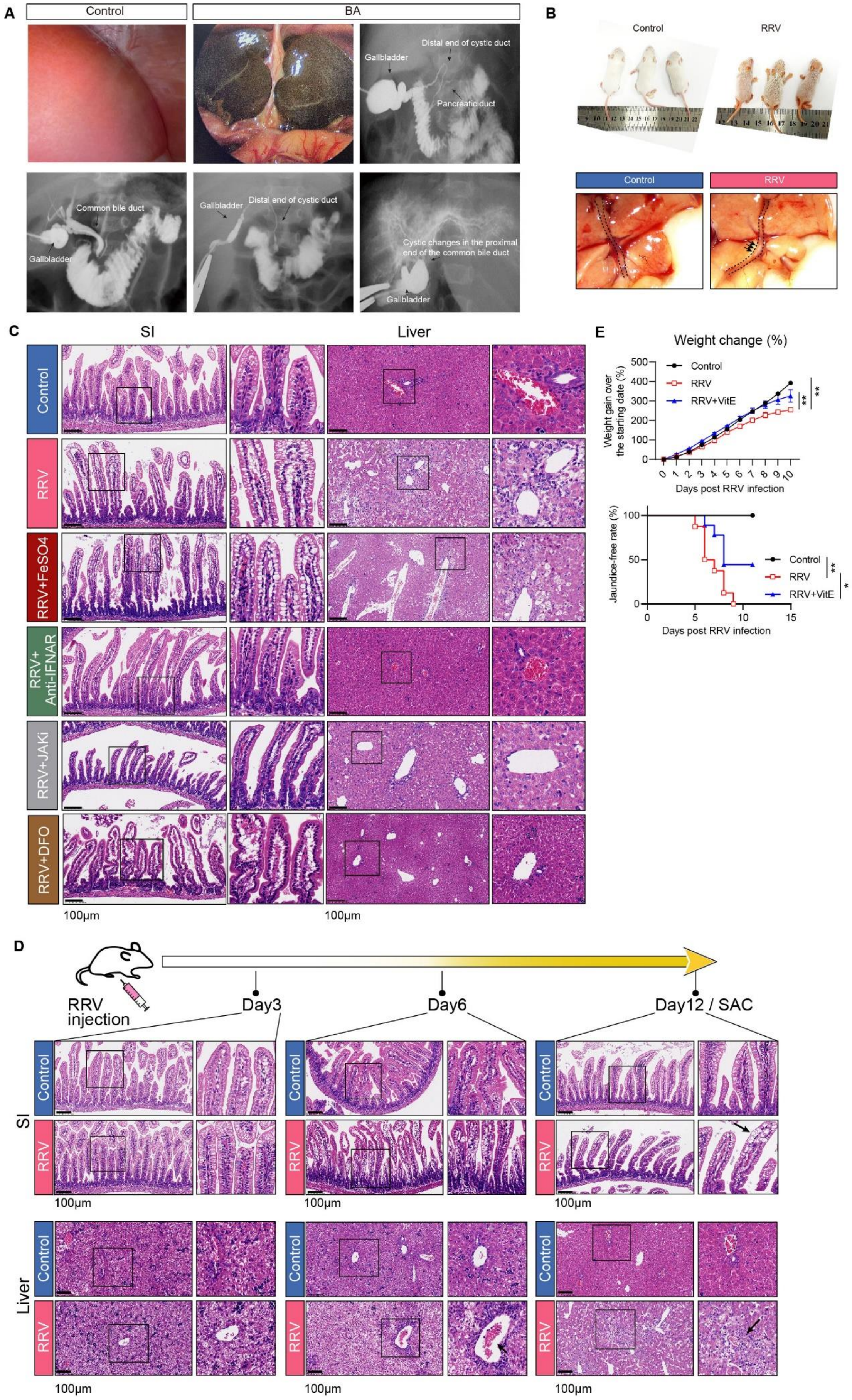

A
Control
BA
Distal end of cystic duct
Gallbladder
Pancreatic duct
Common bile duct
Gallbladder
Gallbladder
Distal end of cystic duct
Cystic changes in the proximal end of the common bile duct
Gallbladder
B
Control
RRV
Control
RRV
C
SI
Liver
Control
RRV
RRV+FeSO4
RRV+ Anti-IFNAR
RRV+JAKi
RRV+DFO
100μm
100μm
E
Weight change (%)
Weight gain over the starting date (%)
Control
RRV
RRV+VitE
Days post RRV infection
Jaundice-free rate (%)
Control
RRV
RRV+VitE
Days post RRV infection
D
RRV injection
Day3
Day6
Day12 / SAC
SI
Liver
Control
RRV
100μm


59

**Supplementary Figure 5. IFN-I-JAK-STAT signaling contributes to BA pathology.** (**A**) Typical BA appearance under laparoscopy and cholangiography for the patients. (**B**) Typical BA appearance in RRV-infected mice under stereoscope was shown. (**C**) Representative hematoxylin-eosin (H&E) stained sections for liver and small intestine biopsies of control, RRV and RRV-infected mice with indicated treatments. (**D**) Representative H&E stained sections for liver and small intestine biopsies of control and BA mice at the $3^{rd}$, $6^{th}$ and $12^{th}$ day post RRV infection. (**E**) Line charts displaying body weight changes (upper) and jaundice occurrence rates (lower) in control and RRV-infected mice with VitE treatment. The statistics for body weight change was performed using two-way ANOVA, for jaundice-free survival rate was performed using Log-rank test; *$p < 0.05$, **$p < 0.01$, ***$p < 0.001$.

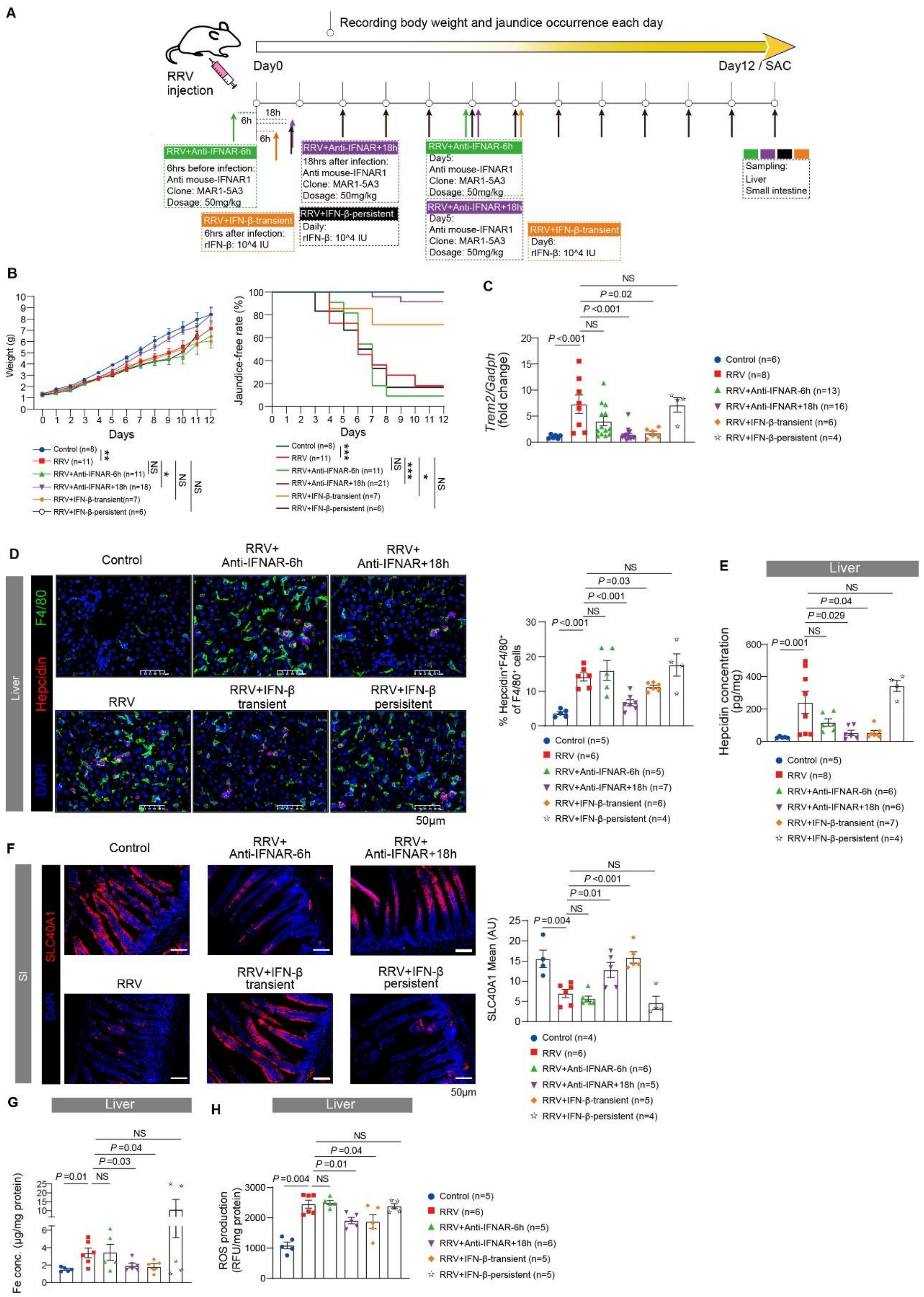


**Supplementary Figure 6. Transient but not persistent IFN-I activation improves BA outcomes in RRV-infected neonatal mice.** (**A**) Schematic of treatment strategy. Body weight and jaundice occurrence were recorded each day. At 12 days post RRV infection, mice were sacrificed for sampling. (**B**) Line charts displaying body weight changes and jaundice occurrence rates. The statistics for body weight change was performed using Two-Way ANOVA, for jaundice-free survival rate and survival probability were performed using Log-Rank test; *$p < 0.05$, **$p < 0.01$, ***$p < 0.001$. (**C**) Relative expression of *Trem2* in liver were examined by qPCR. (**D**) Representative immunofluorescent images showing co-staining of F4/80 (green) and hepcidin (red) in liver. Scale bar, 100μm. Dot plots showing percentage of F4/80$^{+}$hepcidin$^{+}$ cells of F4/80$^{+}$ cells in the liver biopsies. (**E**) Dot plots showing concentrations of hepcidin in liver homogenates from control (n=5), RRV-infected mice (n=8) and with indicated treatment (RRV+anti-IFNAR-6hrs, n=6; RRV+anti-IFNAR-18hrs, n=6; RRV+IFN-β-transient, n=7; RRV+IFN-β-persistent, n=4). Hepcidin concentration measured as per mg protein. *P* values were calculated by two-tailed Student's t test. (**F**) Representative immunofluorescent images showing SLC40A1 staining in the small intestine biopsies from control (n=5), RRV-infected mice (n=6) and with indicated treatment (RRV+anti-IFNAR-6hrs, n=5; RRV+anti-IFNAR-18hrs, n=5; RRV+IFN-β-transient, n=5; RRV+IFN-β-persistent, n=4). Scale bar, 50μm. Measurement of mean fluorescence intensity (MFI) for SLC40A1 and the statistical unit was shown as Arbitrary Unit (AU). (G) Dot plots showing concentrations of iron in liver homogenates from control (n=4), RRV-infected mice (n=6) and with indicated treatment (RRV+anti-IFNAR-6hrs, n=6; RRV+anti-IFNAR-18hrs, n=6; RRV+IFN-β-transient, n=5; RRV+IFN-β-persistent, n=5). Iron concentration was expressed as per mg protein. (H) Dot plots showing hepatic ROS production from control (n=5), RRV-infected mice (n=6) and with indicated treatment (RRV+anti-IFNAR-6hrs, n=5; RRV+anti-IFNAR-18hrs, n=5; RRV+IFN-β-transient, n=5; RRV+IFN-β-persistent, n=5). ROS concentration was expressed as per mg protein. P values were calculated by two-tailed Student's t test for all experiments.

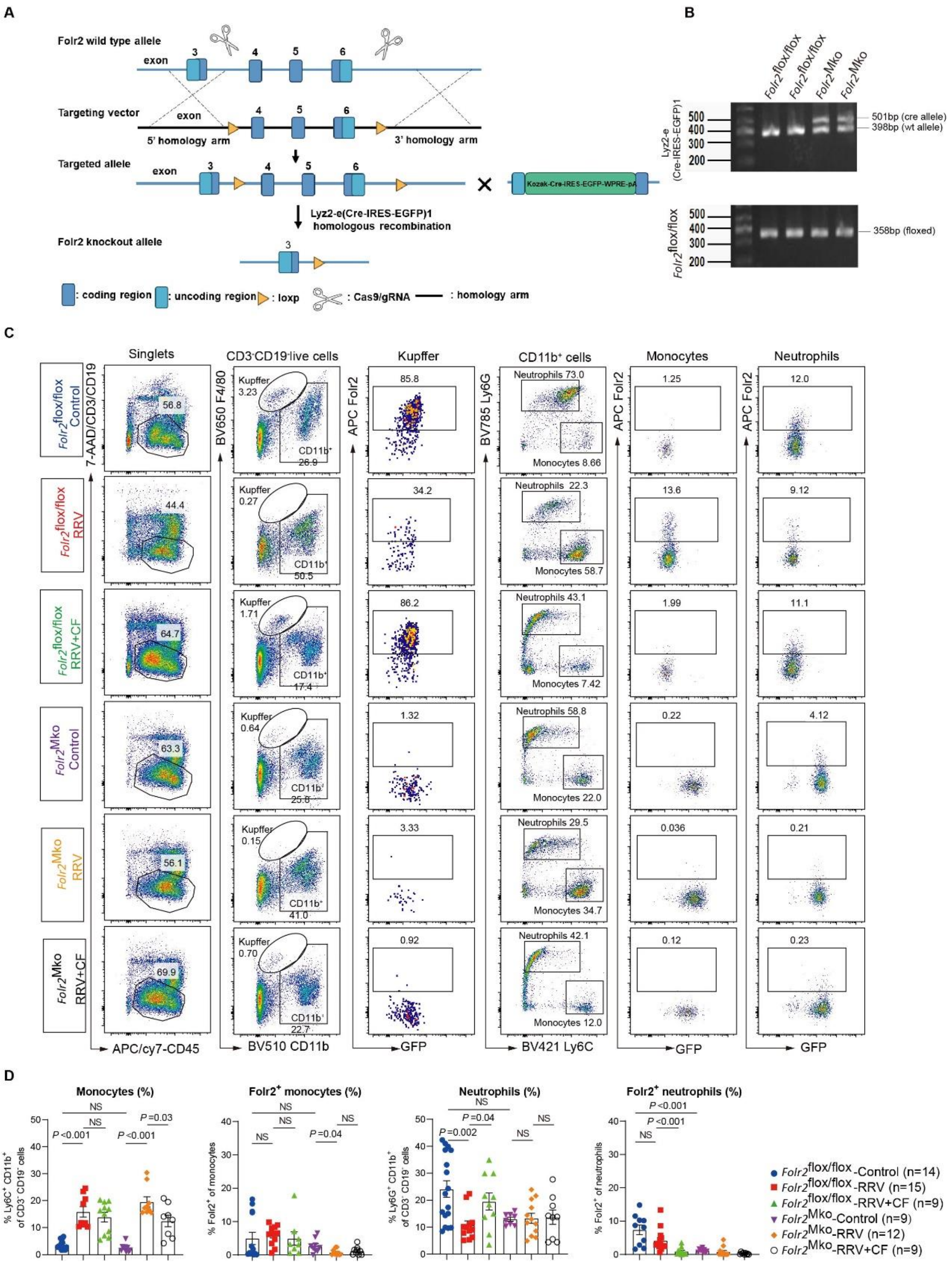


**Supplementary Figure 7. Establishment of myeloid-specific *Folr2* knockout mice (*folr2$^{Mko}$*) on BALB/C background.** **(A)** Strategies for establishment of myeloid-specific *Folr2* knockout mice (*Folr2*$^{\mathrm{Mko}}$). **(B)** Gel electrophoresis showing representative genotyping data for *Folr2$^{Mko}$* and *Folr2$^{flox/flox}$* mice. **(C and D)** Gating scheme for comparing FOLR2 expression in Kupffer cells (CD11b$^{int}$F4/80$^{+}$), monocytes (CD11b$^{+}$Ly6C$^{hi}$) and neutrophils (CD11b$^{+}$Ly6G$^{hi}$) in the livers of *Folr2$^{Mko}$* mice compared with *Folr2$^{flox/flox}$* mice under control, RRV and RRV+CF treatment. *P* values were calculated by two-tailed Student's t test.

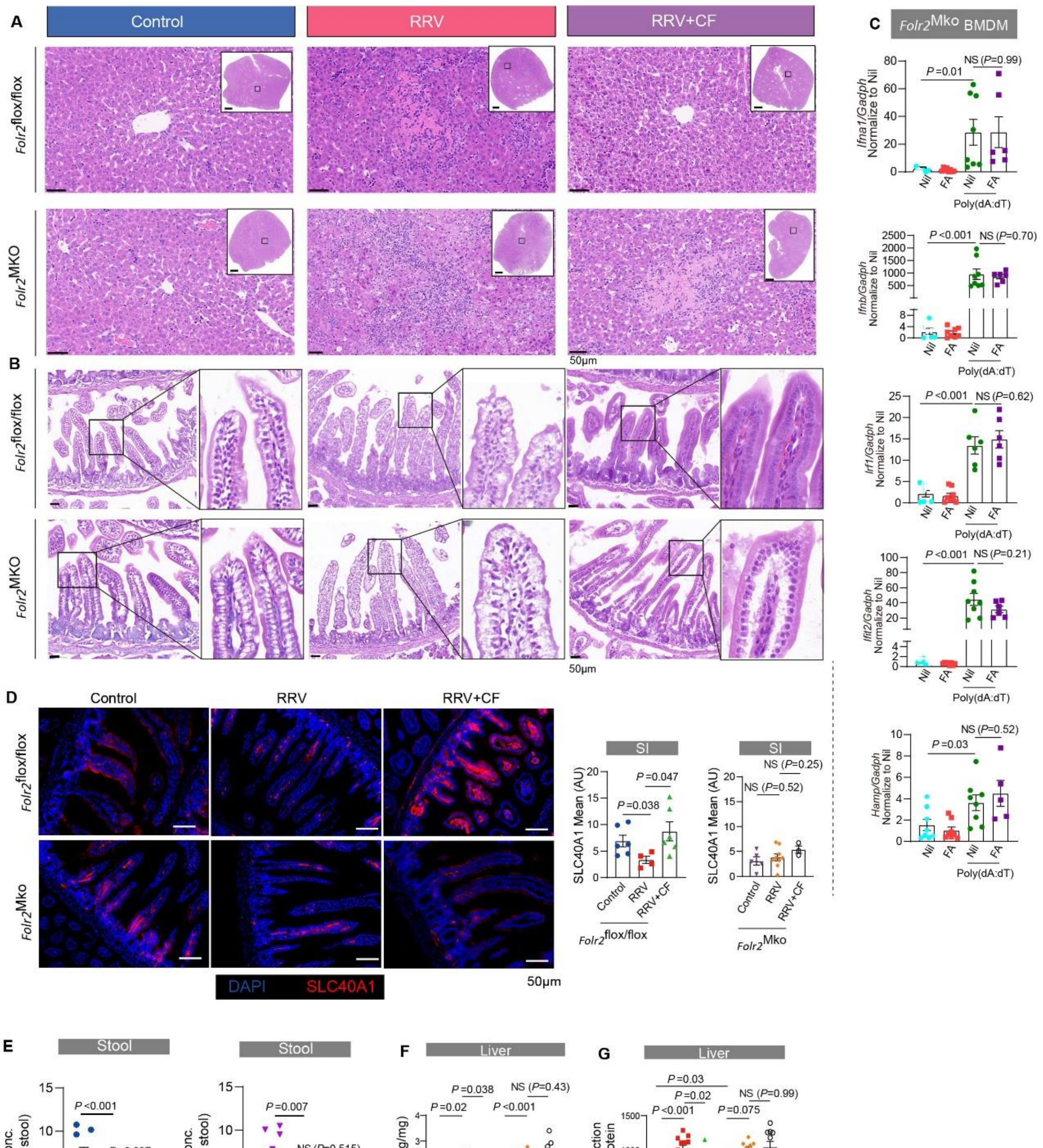


**Supplementary Figure 8. Loss of therapeutic effects of calcium folinate in *Folr2*$^{Mko}$ mice. (A and B)** Representative H&E sections for liver and small intestine biopsies of *Folr2*$^{MKO}$ mice compared with *Folr2*$^{flox/flox}$ mice under control, RRV and RRV+CF treatment. (**C**) Relative expression of *Ifna1*, *Ifnb*, *Irf1*, *Ifit2, and Hamp* were examined by qPCR in BMDM from *Folr2*$^{MKO}$ mice transfected with Poly (dA: dT) with or without folic acid (FA) treatment. Data were normalized to the relative expression of indicated genes in Nil (unstimulated) conditions and are presented as means ± SEM. *P* values were calculated by two-tailed Student's t test. (D) Representative immunofluorescent images showing SLC40A1 (red) in small intestine from control and RRV-infected

Folr2flox/flox and Folr2Mko mice with or without CF treatment. Scale bar, 50μm. Measurement of mean fluorescence intensity (MFI) for SLC40A1 and the statistical unit was shown as Arbitrary Unit (AU). (**E**) Dot plots showing iron content in stool from control and RRV-infected *Folr2*$^{flox/flox}$ and *Folr2*$^{Mko}$ mice with or without CF treatment. Iron concentration was expressed as per mg weight. Line represents the mean ± SEM. (F) Dot plots showing concentrations of iron in liver homogenates from control, RRV-infected *Folr2*$^{flox/flox}$ and *Folr2*$^{Mko}$ mice and with CF treatment. Iron concentration was expressed as per mg protein. (G) Hepatic ROS production measured and expressed as per mg protein. *P* values were calculated by two-tailed Student's t test for all experiments.

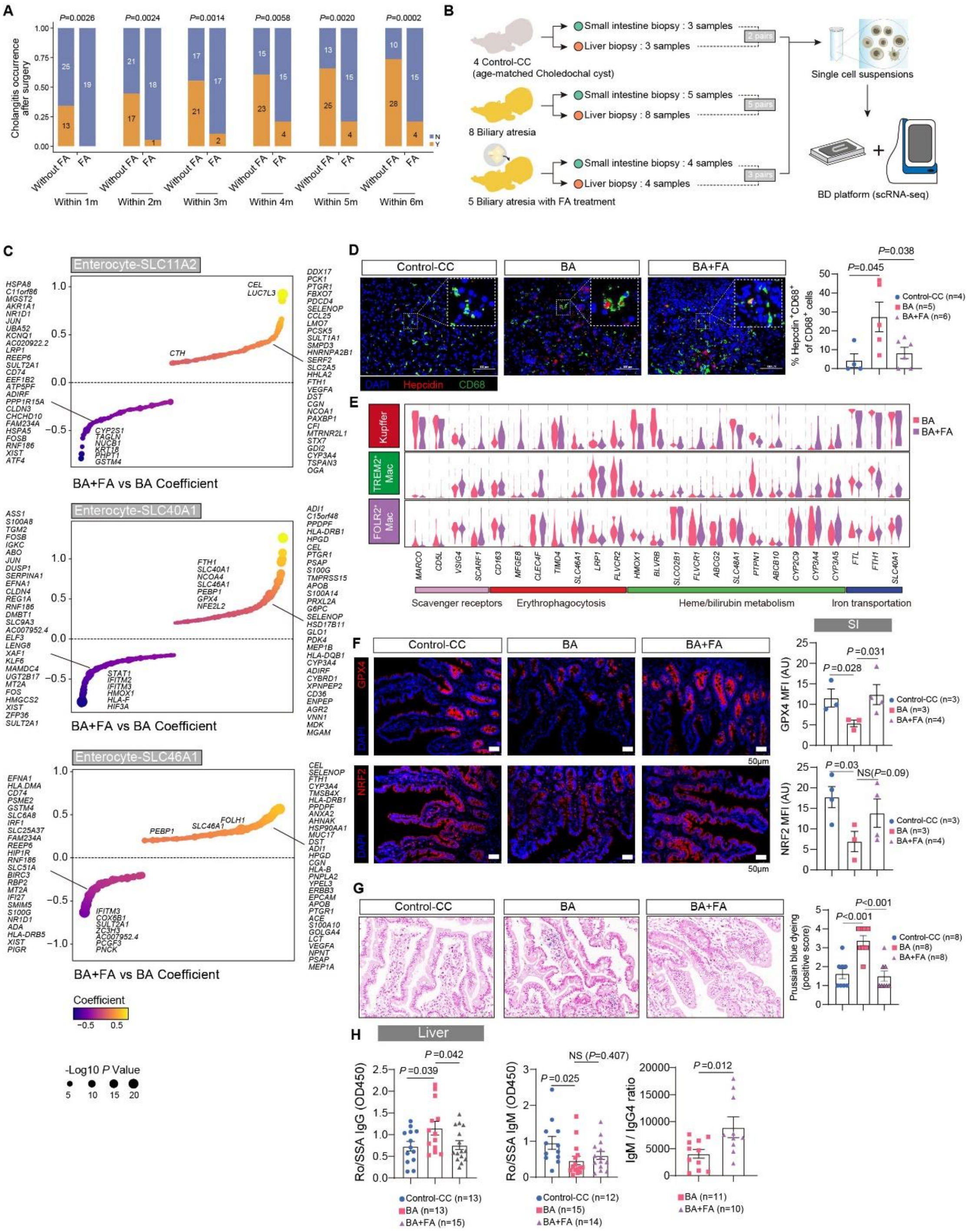


**Supplementary Figure 9. An open label clinical study of folic acid in infants with BA demonstrate therapeutic effects.** (**A**) The monthly record of postoperative cholangitis occurrence was shown in stacked bar plots within 6-month follow-up of the patients with or without FA treatment. The number of patients with different outcomes in two groups are labeled. *P* value was determined by Chi-Squared test. (**B**) Schematic diagram of the experimental design for single cell sequencing based on BD Rhapsody$^{TM}$ platform. (**C**) Scatter plots showing differentially expressed genes of enterocyte-SLC11A2, enterocyte-SLC40A1 and enterocyte-SLC46A1 cells between patients with (BA+FA) and without FA supplement (BA). Colored dots denote the up-regulated and down-regulated genes passing the thresholds of P value <=0.05 and |Coefficient| >=0.25, respectively. Dot color and size are scaled by the coefficient and *P* value, respectively. (D) Representative immunofluorescent images showing co-staining of CD68 (green) and hepcidin (red) in liver from control-CC (n=4), BA (n=5) and BA with FA treatment (n=4). Scale bar, 100μm. Dot plots showing comparison of percentage of $CD68^{+}hepcidin^{+}$ cell in total $CD68^{+}$ cells. Each point represents an individual patient, and the line represents the mean ± SEM. *P* values were calculated by two-tailed Student's t test. (**E**) Violin plots showing comparison of genes involving scavenger function, erythrophagocytosis, heme/bilirubin metabolism and iron transportation in hepatic Kupffer cells, $TREM2^{+}$Mac and intestinal $FOLR2^{+}$Mac between BA and BA+FA subjects. (**F**) Representative immunofluorescent images showing GPX4 and NRF2 staining in small intestine biopsies from control-CC, BA and BA+FA subjects. Scale bar, 50μm. Measurement of mean fluorescence intensity (MFI) for NRF2 and GPX4 and the statistical unit was shown as Arbitrary Unit (AU). P values were calculated by one-tailed Student's t test. (**G**) Representative Prussian blue staining images showing ferric ($Fe^{3+}$) iron in small intestine biopsies from control-CC, BA and BA+FA subjects. Scale bar, 50μm. The statistical unit was shown as "positive score". According to the intensity of Prussian blue staining, "0" shows no blue color; "1+" has a small amount of iron particles or occasional small amounts of iron beads; "2+" has a large number of iron particles and small iron beads; "3+" has many iron particles, small beads, and a few small blue and black pieces; "4+" has a lot of iron particles and small beads, and many densely packed small pieces. *P* values were calculated by two-tailed Student's t test. (**H**) Dot plots displaying the concentrations of anti-Ro/SSA IgG (left) and IgM (right), and ratio of IgM to IgG4 in liver biopsies from control, BA and BA+FA groups. P values were calculated by two-tailed Student's t test.

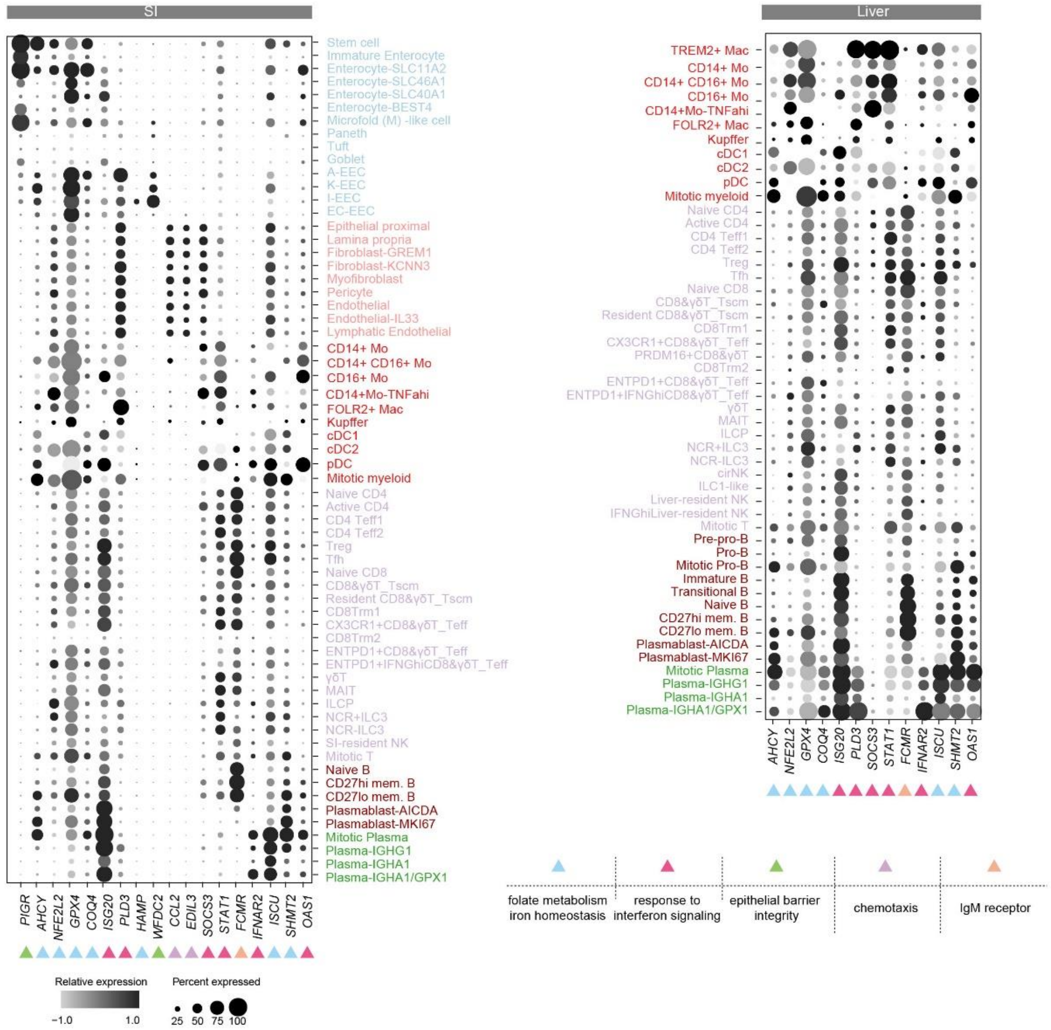


**Supplementary Figure 10. A genome-wide association study (GWAS) for infants with BA identify candidate risk genes.** Dot plots showing the expressions of selected candidate risk genes in all functional subsets from small intestine (left) and liver (right). The enriched biological processes for the selected risk genes are marked by different color arrows and depicted within the chart.
