## Supplementary materials and methods for "Folic acid prevents interferon-induced iron accumulation and ferroptosis and improves liver health in children with biliary atresia"

#### Mouse models of influenza infection

Female sucking BALB/C mice at 14 days of age were raised with the mother mice in microisolator cages at a BSL-2 laboratory at Guangzhou Medical University, under standard laboratory conditions (room temperature 22°C ± 2°C, relative humidity 50% ± 10%) and a 12h/12h light/dark cycle. Housing and handling of mice was in accord with the Institutional Animal Care and Use Committee of Guangzhou Medical University on Animal Care guidelines. Influenza virus A (H1N1; PR8) was used. PR8 virus at 1000 plaque-forming units (PFU) in 50 µL of PBS or PBS only (control) were intranasally inoculated into the mice under inhalation anesthesia with isoflurane. Body weight was monitored daily after infection, and mice were euthanized when weight loss reached 20%. Their liver, small intestine and blood samples were collected.

#### Mouse models of bile duct ligation (BDL)

6-week-old male BALB/C mice were purchased from Beijing Vital River Laboratory Animal Technology Co. Ltd. Mice were randomly allocated to BDL and sham groups. Both procedures were performed under sterile conditions. In BDL group, the common bile duct was isolated from the flanking portal vein, doubly ligated with silk 5-0 and cut in between ligatures. The common bile duct was isolated without ligation in sham group. They were housed with 12h/12h light and dark cycles and with access to food and water in conventional conditions. Body weight and jaundice occurrence were daily recorded. Mice were sacrificed 7 days post-surgery.

#### Iron/ferritin concentration detection

Iron in stool and tissue was measured by plasma and tissue iron concentration detection kit (Solarbio, Cat#BC1735& BC4355) respectively according to the protocols provided by the manufacturer. Briefly, tissues were ground in cold lysis buffer and supernatants after centrifugation were collected for subsequent procedures. Standards and blank controls were set up as instructed. Samples, reagent I and reagent II were mixed thoroughly in 1.5ml Eppendorf tubes and incubated in boiling water bath for 5min. Chloroform was added and mixed thoroughly, and 200µL supernatant was collected after centrifugation. The absorbance at the 520 nm was measured with Multiskan™ FC microplate tester (Thermo Scientific). Iron concentration of liver and small intestine were expressed as per mg protein, which were estimated from BCA protein concentration using Thermo Pierce™ BCA Protein Assay Kits (Thermo Scientific™, Cat#23225). Iron concentrations of stool samples were normalized to per mg weight. Measurement of iron and ferritin in plasma/serum were conducted by department of clinical laboratory at GWCMC. Iron contents were quantitatively determined by using commercialized Iron Ferene Reagent Kit provided by Zhongyuan Huiji Biotechnology Co., Ltd. Ferritin was determined by Direct Chemiluminescence Method provided by Siemens Healthcare Diagnostics Inc.

**Non-heme iron assay**

Equal weight of tissues were homogenized in high-purity water 1:10 (w/v). Protein in homogenates was precipitated with concentrated hydrochloric acid and trichloroacetic acid at 95°C for 1h, then centrifuged at 16,000 rpm for 10min. Supernatant aliquots were mixed with chromogenic solution. Sample blanks were set and the ferrozine chromogenic agent was added to the blank tube. Absorbance at 562nm was measured with Multiskan™ FC microplate tester (Thermo Scientific™). Iron concentration of liver and small intestine were expressed as per mg protein, which were estimated from BCA protein concentration using Thermo Pierce™ BCA Protein Assay Kits (Thermo Scientific™, Cat#23225).

**Lipid peroxidation (4-HNE) assay**

Tissue 4-HNE was determined using 4-HNE Assay Kit (Abcam, Cat#ab238538). Liver and small intestine samples were homogenized with PBS with 0.1% BSA on ice. ELISA plates were coated with 10μg/ml 4-HNE conjugate overnight at 4°C. After blocking, 50μL sample homogenates, standards (diluted according to the instructions) and 50ul Anti-4HNE detection antibody were added, followed by incubation with Secondary Antibody-HRP Conjugate at room temperature for 1h. The enzymatic activity was detected with substrate solution. The reaction was stopped and measured at 450nm wavelengths on a microplate reader. 4-HNE concentration of liver and small intestine were expressed as per mg protein estimated from BCA protein concentration in respective samples using Thermo Pierce™ BCA Protein Assay Kits.

**ROS/RNS detection**

DCF ROS/RNS Assay Kit purchased from Abcam (Cat#ab238535). Specific ROS/RNS fluorescence detection probe (dichlorodihydrofluorescin DiOxyQ) firstly underwent quenching removal and then was diluted in 1X stabilizing solution. Equal amount of tissue homogenates, catalyst and DCFH solution were mixed for rapid reaction. The relative fluorescence unit (RFU) was measured at 480nm excitation and 530nm emission. Tissue homogenates were subjected for protein quantification by BCA, and the final normalized results were expressed as RFU per mg protein.

**Enzyme-linked immunosorbent assay (ELISA)**

Detection of hepcidin (Cat#SEB979Bo), folic acid (Cat#CEA610Ge), glutathione (GSH) (Cat#CEA294Ge), homocysteine (HCY) (Cat#PAD984Ge01), and tetrahydrobiopterin (BH4) (Cat# CPG421Ge21) was accomplished with ELISA kit as per the manufacturer’s instructions (Cloud-Clone Corp, Wuhan, China). Pre-experiments were performed to ensure optimized dilution of samples and incubation conditions. Total protein in liver and small intestine were determined by BCA protein concentration in respective samples using Thermo Pierce™ BCA Protein Assay Kits.

**Detection of HCMV/RV human antibodies**

Serum levels of IgM-, IgG- and IgA-specific antibodies targeting human rotavirus (RV) and human CMV (HCMV) in age matched control-healthy, control-CC, BA and

HCMV$^{+}$ NH were measured by ELISA purchased from Jiangsu Meimian Industrial Co., Ltd. Pre-experiments were performed to ensure optimized dilution of serum and incubation conditions.

**Murine interferon-beta (mIFN-β) bioluminescent ELISA**

LumiKine™ Xpress mIFN-β 2.0 is an 'sandwich' ELISA kit for mIFN-β detection (InvivoGen, Cat# luex-mifnbv2). White flat-bottom MaxiSorp® 96-well plate (Thermo Scientific™, Cat# 437591) was coated overnight with 50 µl mIFN-β capture antibody at room temperature. After blocking, 50ul of liver homogenates, standards, and 30 ng/ml lucia-conjugated detection antibody were added and incubated at 37℃ for 2h. After washing, 50ul reconstituted QUANTI-Luc™ Plus was added and immediately detected by chemiluminescence. The microplate reader was set in advance, and the reading time was set to 0.5s.

**Multiplex assays of cytokines**

Bio-Plex Pro™ Human Inflammation Panel 1, 37-Plex was used according to protocols provided by the manufacturer (Bio-Rad Laboratory, Cat#171AL001M). Briefly, filter plate was pre-wet and magnet beads was added. After washing, samples with appropriate dilution and standards were added and incubated at room temperature at 850rpm on Eppendorf ThermoMixer® C (Eppendorf, Cat#2231001005) for 30 minutes, followed by incubation with detection beads on shaker for 30 minutes. SA-PE was then added and incubated for 10 minutes. Data were collected and analyzed using a Bio-Rad Bio-Plex 200 instrument equipped with Bio-Plex Manager software version 6.0 (Bio-Rad Laboratory). The precision based on both intra and inter-assays variations were < 10% within the detection limits provided by the manufacturer. The immunoassay data in liver and small intestine were expressed as per mg protein estimated from protein concentration in respective tissue homogenates samples using Thermo Pierce™ BCA Protein Assay Kits.

**Transfection of Bone Marrow Derived Macrophage (BMDM)**

Primary bone marrow cells, which were obtained from BALB/C suckling mice (WT, *Folr2*$^{flox/flox}$ and *Folr2*$^{Mko}$), were cultured in RPMI1640 medium (Gibco™, Cat#11875093) supplemented with 10% FBS, 1% penicillin/streptomycin and M-CSF (50ng/ml) (PEPROTECH, Cat#300-25) for 5 days. The adherent cells were bone marrow monocyte-derived primary macrophages (BMDM). Time-dependent Folr2 expression was measured by flow cytometry. BMDMs were subjected to Poly(dA:dT) (5ug/ml) (InvivoGen, Cat#tlrl-patn-1) transfection using Superluminal High-efficiency Transfection Reagent (InvivoGen) with or without folic acid (10ng/ml) (Merck, Cat# F7876) stimulation for 18 hours. Cell lysates were collected for RNA extraction and RT-qPCR analysis.

**Immunofluorescence staining/multiplex staining**

Paraffin-embedded sections of liver and small intestine biopsies from patients and mouse models were processed. Briefly, paraffin sections were placed at a 65℃ drying oven for 1h, and transferred into xylene for dewaxing and hydrated in 100%, 90%, 80%, 70% ethanol successively. Antigen retrieval was performed in Citrate Buffer or TE buffer (pH6.0 or pH9.0, accordingly to primary antibody requirements) with microwave treatments (high-power for 5 min, and low-power for 10 min). Followed by blocking with 10% goat serum with 0.3% Triton-X100 for 2h at room temperature (RT), slides were incubated with primary antibodies in a humidified chamber at 4°C overnight. After washing, secondary antibody was incubated for 1 hour at room temperature. Spontaneous fluorescence quenching was then performed with the TrueVIEW™ Autofluorescence Quenching Kits (Vector, Cat# SP-8400-15) for 10min at room temperature.

For multi-staining, opal polymer HRP Ms+Rb secondary antibody was added for 10 min at RT, TSA (Tyramide signal amplification) visualization was performed with OPAL 7-Color Manual IHC Kit (Akoya biosciences, Cat# OP-000003) containing the following fluorophores: Opal 520, Opal 540, Opal 570, Opal 620, Opal 650 and Opal 690. Slides were incubated for 10 min at RT in the dark, followed by washes with TBST. Multiplex were performed by repeating staining cycles in series, with microwave treatments between each step to remove the antibody complex. For the last step, slides were counterstained with VECTASHIELD Antifade Mounting Medium with DAPI (Vector, Cat#H-1200-10) for nuclear staining. Immunofluorescent images were acquired with Leica DMi8 Inverted Fluorescence Microscope (Leica Microsystems) and Olympus BX53 biological microscope, Panoramic fluorescence scanning was performed with KF-SCI-005 Digital Section Fluorescence Scanner (Jiangfeng, China) and Vectra Polaris Multi-color Fluorescence Quantitative Analyzer (Akoya Biosciences). The relative fluorescence intensity was calculated by ImageJ software. Information of all primary and secondary antibodies used in Immunofluorescence staining/multiplex staining were summarized in Supplementary table 6.

**Immunohistochemistry**

Processing procedures of paraffin sections were consistent with that of immunofluorescence staining. Afterwards, slides were incubated with Goat anti-rabbit/mouse-HRP IgG the secondary antibody 30 minutes at room temperature. Subsequently, the peroxidase label was visualized with Pierce™ DAB Substrate Kit (Thermo Scientific™, Cat#34002). Slides were then counterstained with hematoxylin (Merck, Cat#MHS16) for 1 minute, rinsed with distilled water, dehydrated, and cover slipped. The proportion of positive cells was analyzed by ImageJ software, and the results are shown as the percentage of positive cells.

**Transmission electron microscopy (TEM)**

Liver and small intestine biopsies obtained from surgery operation were immediately fixed in 2.5% glutaraldehyde solution and stored at 4°C overnight. After three washes in PBS, tissues were fixed in 1% osmium acid for 2h, then dehydrated in graded ethyl alcohol series and embedded in SPI-Pon 812 epoxy resin (SPI) overnight at 37°C. The polymerization was performed at 60°C for 48 h. Ultrathin sections (60–80 nm) were cut using a Leica UC7 ultramicrotome (Leica Microsystems), stained with 2% uranyl acetate and lead citrate for 15 min, and viewed under an HT7700 transmission electron microscope (TEM, HITACHI).

**Hematoxylin and Eosin (H&E) staining**

Fresh small intestine and liver tissues were fixed in 4% paraformaldehyde. Small intestine tissues from mouse models were wrapped into Swiss roll and cut longitudinally. Tissues were dehydrated, cleared and embedded in paraffin, cut into serial 4 μm thick slices and stained with H&E for histological analysis.

**Prussian blue iron stain**

Paraffin sections were dewaxed by xylene and hydrated with gradient ethanol. Prepare Lillie $Fe^{3+}$ Dyeing Solution (Liquid A (potassium ferricyanide) mixed with liquid B (dilute acid in equal proportions) according to manufacturer's guidelines. Paraffin sections were stained with Dyeing Solution in dark for 30mins, and washed with distilled water for 5mins. Nuclear staining was carried out by NuclearFastRedStaining Solution for 10min, followed by rinsing with distilled water for 2min. After conventional dehydration with graded ethanol and xylene, slides were dried and sealed with neutral balsam. According to the intensity of Prussian blue staining, "0" shows no blue color; "1+" has a small amount of iron particles or occasional small amounts of iron beads; "2+" has a large number of iron particles and small iron beads; "3+" has many iron particles, small beads, and a few small blue and black pieces; "4+" has a lot of iron particles and small beads, and many densely packed small pieces.

**Flow cytometry**

Liver and small intestine was digested to single cell suspensions. To distinguish subtypes of granulocytes, we stained cells with 12-color combination of the following monoclonal antibodies: CD45, CD16, CD14, CD11b, CD68, TIM-4 and FOLR2. All antibodies were purchased from Biolegend. Information of antibodies were summarized in table S6. Stained cells were analyzed with a BD Fortessa X20 (BD biosciences). Prior to flow cytometric acquisition, 7-AAD was added for exclusion of dead cells. For detection of IFN-γ expression in hepatic T cells, liver single cell suspensions were stimulated with PMA (100 ng/mL), ionomycin (1 ng/mL) and monensin (2 mM) for 4 hours. Cells were surface stained with anti-CD45, CD4, CD3, CD8, after fixation and permeabilization, cells were intracellularly stained with anti-IFN-γ and IL-17A antibodies. IFN-γ expression was analyzed in hepatic T cells. Zombie dyes was added to exclude dead cells. Data were analyzed with FlowJo.

**RNA extraction and RT-qPCR**

The phenol-chloroform based RNA extraction from cells and tissues was performed. cDNA was reverse transcribed using HiScript III Reverse Transcriptase (Vazyme, Cat#R302) according to the manufacturer's protocol. RT-qPCR was performed with HiScriptII One Step QRT-PCR SYBR Green Kit (Vazyme, Cat# Q221) using the ABI

7500 Fast system (Applied Biosystems). The primers are listed in table S5. Expression levels were determined with triplicate assays per sample.

**RNA-seq library construction for 10x Genomics single cell 5' and V(D)J sequencing**

Liver and small intestine biopsies were processed as described [1, 2]. For liver, $CD45^+$ live cells were flow-sorted by a FACSAria SORP flow cytometer (BD Sciences). For small intestine, single cell suspensions were obtained after enzymatic digestion and dead cell removal (Miltenyi Biotec). Single-cell 5' RNA sequencing libraries were constructed according to the protocols of the Chromium Single Cell 5' Library kit (10x GENOMICS). In brief, single-cell suspension was mixed with RT-PCR master mix and loaded onto the 5' chip. Then, RNA transcripts were uniquely barcoded within each cell and reverse-transcribed into barcoded cDNA, followed by purification, amplification and adaptor ligation. TCR- and BCR- enriched libraries were generated with aliquots from each of the aforementioned cDNA using the Chromium Single Cell V(D)J Enrichment kit. All libraries were sequenced on the Illumina Novaseq 6000 platform.

**RNA-seq library construction for BD Rhapsody™ single-cell analysis system**

Liver and small intestine biopsies from age matched infants (8 BA, 4 age-matched control, 5 BA with FA treatment) were processed as described above. BD Rhapsody™ Human Single-Cell Multiplexing Kit (BD biosciences) was utilized for scRNA library construction. Up to 2 samples were labelled and pooled prior to single cell capture with the BD Rhapsody™ Single-Cell Analysis system. After partitioning and lysis of cells, cDNA is encoded on BD Rhapsody™ Enhanced Cell Capture beads using both the 3' and 5' ends of transcripts as templates. Whole transcriptome mRNA libraries are amplified using random priming of the on-bead cDNA libraries.

**Preprocessing of single-cell RNA-seq data**

The raw sequence reads were demultiplexed and aligned to the human transcriptome reference (GRCh38) using CellRanger-3.1.0 (for 10X Genomics) or a cwl pipeline (for BD Rhapsody™ system) (https://bitbucket.org/CRSwDev/cwl/src/master/). The raw expression count matrix (gene counts versus cells) was generated for each sample and transformed to Seurat object using the R package Seurat v4.0.0 [3]. Qualified cells are identified as cells with 200-5000 detected genes and with less than 50,000 detected unique molecular identifiers (UMIs). Following quality control, the filtered data objects of all samples were merged and normalized using the Seurat's SCTransform function to eliminate the impacts of sequencing depth, mitochondrial gene expression and other batch effects. A total of 242,327 high quality cells generated using 10X Genomics platform and 210,661 cells generated using BD Rhapsody™ system were included in subsequent analysis.

**Identification of major cell types and functional cellular subsets**

The normalized gene expression matrix was subjected to dimension reduction and clustering to identify major cell types. Variably expressed genes were selected for principal component analysis (PCA). The first 30 principal components were used for uniform manifold approximation and projection (UMAP) dimension reduction, shared-nearest neighbor computation and cluster identification. The identified clusters were allocated to major cell types by mapping canonical cell markers in the two-dimensional UMAP map, and a total of 7 major cell types were identified for the small intestine and liver.

We then analyzed functional sub-clusters within each of major cell types. PCA was performed using the variably expressed genes for each major cell type object by using the “SCT” assay mode. Significant principal components were used for UMAP dimension reduction and clustering. Seurat function FindClusters was utilized with adapted resolution to identify cellular subsets within the 7 major cell types. Of note, cells expressing dual-lineage markers were removed from downstream analysis to reduce the impact of potential doublet capture bias. Differentially expressed genes (DEGs) of each cellular subset were identified using Seurat function FindAllMarkers. Detailed descriptions of each cellular subset and the marker genes are shown in the figures and main text of the relevant sections.

**Identification of differentially expressed genes between control and disease or treatment versus non-treatment**

The identification of differentially expressed genes (DEGs) between control and disease of each functional cellular subset was performed using MAST[4], which fits a hurdle model for handling zero-inflated single cell assay data and comparing continuous gene expression level by using linear regression. Specifically, we applied the following regression model formula to analyze the effects of groups (disease versus control or treatment versus non-treatment group) and tissue origins (small intestine versus liver) on gene expression,

$$\mathrm{Yi} \sim \mathrm{G} + \mathrm{N} + \mathrm{T}$$

in which, Yi represents the gene expression value for gene i across all cells (by using the “RNA” assay mode of each Seurat object), G represents the group to be compared with the reference, and N represents the total number of detected genes in each cell (to control the complexity for different cells). The coefficients for each variable in the regression model were retrieved and calculated the significance using the likelihood ratio test in MAST. We set the thresholds of P value <=0.05 and |Coefficient| >=0.25 to identify up- or down-regulated genes between the given different groups.

**Identification of transcriptional signature and pathway analysis**

For the functional analysis of cellular subsets within each major cell type, we performed oposSOM[5], which is based on a self-organizing map (SOM) machine learning strategy, to retrieve the transcriptional signatures. For the lineage of interest (e.g., myeloid cells), the expression matrix was used as input to construct a self-organizing map and identify co-expressed metagene modules. After identification of metagenes and signature, we calculated the average expressions of each metagene in each cell. By applying the following threshold criteria,

$$\mathrm{e}^{\mathrm{threshold}} = 0.5 \times \mathrm{e}^{\mathrm{max}}$$

we defined the significant expressions of different metagenes in each cell.

Functional enrichment analysis for DEGs or metagenes was conducted using Metascape[6] or GSVA[7]. GSVA identifies priori-defined gene sets showing significant different expressions between two given groups. The enriched pathways were based on Gene Ontology biological process terms and KEGG pathways.

**Analysis of transcription factor activity**

The gene regulatory networks across different cellular subsets were predicted using SCENIC[8]. In brief, the co-expression network of genes with specific transcriptional factors (TFs) were first identified using the function runGenie3 in the GENIE3 package and converted into co-expression modules by running the function runSCENIC_1_coexNetwork2modules. Then, the potential direct-binding target genes (regulons) for each TF were identified using the function runSCENIC_2_createRegulons implemented in RcisTarget package. Finally, the activities of all the regulons were scored in each cell using runSCENIC_3_scoreCells function implemented in the R package AUCell. The AUCell matrices were retrieved for heatmap plotting.

To better illustrate the regulatory activity between TFs and their target genes, we applied for MAGIC[9] each major cell type to impute the expressing data of all genes and calculated the kNN-DREMI[10] correlation of a given TF-target pair.

**Analysis of 16s rDNA sequencing data**

Paired-end reads from the original DNA fragments were merged and assigned to each sample by the unique barcodes. Sequences analysis were performed using UPARSE package based on the UPARSE-OTU and UPARSE-OTUref algorithms[11]. Sequences with ≥97% similarity were assigned to the same OTUs. The representative sequences for each OTU were selected, and the RDP classifier was used to annotate taxonomic information for each representative sequence. The analysis of diversity, taxonomy and abundance of gut microbial community was conducted using Qiime2[12]. The functional prediction of gut microbial community was analyzed by Picrust2[13]. The difference of microbial abundance and enriched pathways was analyzed using LEfSe[14].

**Genome-wide association study (GWAS)**

Peripheral blood from 297 patients with BA from GWCMC and Liver Transplantation Center, Beijing Friendship Hospital were collected as a case cohort for the GWAS study. The legal guardians of all participants signed informed consent forms for biological investigations. This project was reviewed and approved by the Ethics Committee of GWCMC and Ethics Committee of Beijing Friendship Hospital, in adherence with the Declaration of Helsinki Principles. After quality control, 5,955,093 genotyped or imputed SNPs from 281 cases and 3453 controls were obtained in this study. PLINK (v1.9b) program was used for quality control [15]. Whole-genome imputation of SNPs was carried out by using the IMPUTE2 program [16]. Gene-based association analyses were conducted by using the effective chi-squared test (ECS) method implemented in KGG (v4.0) software [17].

### Genome-wide SNP genotyping and association analysis

The genotyping was conducted by using Infinium Asian Screening Array-24 v1.0 BeadChip arrays (Illumina, Inc., San Diego, CA, USA), according to the manufacturer's protocols. PLINK (v1.9b) program was used for quality control[18]. Samples with genotype call rate < 92%, problematic sex estimation, excessive heterozygosity rate (departure from 3 standard deviations), and cryptic relatedness were excluded. Population stratification was evaluated by principal component analysis (PCA). Low-quality SNPs with genotyping rate < 95%, departure in Hardy-Weinberg equilibrium (HWE, p < 1×10-4), and low minor allele frequency (MAF < 3%) were filtered out. Whole-genome imputation of SNPs was carried out by using the IMPUTE2 program. 1000 Genomes Phase 3 variant set (October 2014, NCBI build b37) was used as the reference panel. Imputed SNPs with INFO < 0.85, accuracy score< 0.9, or MAF < 3% were removed. The SNP associations were tested by the generalized linear model implemented in SNPTEST (v2.5.6) with arguments "-method score -frequentist 1"[19]. Sex and the first 3 PCs were adjusted as covariates. Gene-based association analyses were conducted by using the effective Chi-Squared test (ECS) method implemented in KGG (v4.0) software. Only SNPs located in protein-coding genes and their flanking 5Kb region were considered in gene-based association analyses. Benjamini-Hochberg's method was used to control the false discovery rate (FDR).
